# GWAS of Retinal Vessel Tortuosity Identifies 173 Novel Loci Revealing Genes and Pathways Associated with Vascular Pathomechanics and Cardiometabolic Diseases

**DOI:** 10.1101/2020.06.25.20139725

**Authors:** Mattia Tomasoni, Michael Johannes Beyeler, Sofia Ortin Vela, Ninon Mounier, Eleonora Porcu, Tanguy Corre, Daniel Krefl, Alexander Luke Button, Hana Abouzeid, Konstantinidis Lazaros, Murielle Bochud, Reinier Schlingemann, Ciara Bergin, Sven Bergmann

## Abstract

**Background:** Fundus images allow for non-invasive assessment of the retinal vasculature whose features provide important information on health. Blood vessel tortuosity is a morphological feature associated with many diseases including hypertension.

**Methods:** We analyzed 116 639 fundus images of suitable quality from 63 662 participants from three cohorts, namely the UK Biobank (n = 62 751), SKIPOGH (n = 397), and *OphtalmoLaus* (n = 512). We used a fully automated image processing pipeline to annotate vessels and a deep learning algorithm to determine the vessel type, characterizing these subjects in terms of their median retinal vessel tortuosity specific to arteries and to veins. Tortuosity was measured by the *distance factor* (the length of a vessel segment over its chord length), as well as measures that integrate over vessel curvature. Using these measures as traits, we performed the largest genome-wide association study (GWAS) of vessel tortuosity to date. We assessed gene set enrichment using the novel high-precision statistical method *PascalX*.

**Results:** Higher tortuosity was significantly associated with higher incidence of angina, myocardial infarction, stroke, deep vein thrombosis, and hypertension. We identified 175 significantly associated genetic loci in the UK Biobank; 173 of these were novel and 4 replicated in our second, much smaller, meta-cohort. We estimated heritability at ∼25% using linkage disequilibrium score regression. Vessel type specific GWAS revealed 114 loci for arteries and 63 for veins. Genes with significant association signals included COL4A2, ACTN4, LGALS4, LGALS7, LGALS7B, TNS1, MAP4K1, EIF3K, CAPN12, ECH1, and SYNPO2. These tortuosity genes were overexpressed in arteries and heart muscle and linked to pathways related to the structural properties of the vasculature. We demonstrated that tortuosity loci served pleiotropic functions as cardiometabolic disease variants and risk factors. Concordantly, Mendelian randomization revealed causal effects between tortuosity, BMI and LDL.

**Conclusions:** Several alleles associated with retinal vessel tortuosity point to a common genetic architecture of this trait with cardiovascular diseases and metabolic syndrome. Our results shed new light on the genetics of vascular diseases and their pathomechanisms and highlight how GWASs and heritability can be used to improve phenotype extraction from high-dimensional data, such as images.

**Clinical Perspective:** *What is new?:*

- We automatically estimated arterial and venous tortuosity in over 100k retinal fundus images using image analysis and deep learning.
- GWAS revealed 173 novel loci.
- Mendelian randomization showed that increased venous tortuosity reduces BMI whereas elevated LDL levels reduce the tortuosity of both arteries and veins.
- Measuring tortuosity in terms of the *distance factor*, which is sensitive to total vessel elongation, had higher heritability and more associated loci than other tortuosity measures that are sensitive to local vessel bending.

*What are the clinical implications?:*

- Tortuosity genes were overexpressed in the aorta, tibial artery, coronary artery, and in two heart tissues.
- Higher tortuosity was associated with higher incidence of angina, myocardial infarction, stroke, deep vein thrombosis and hypertension.
- We demonstrated a shared genetic architecture between retinal tortuosity and certain diseases related to the vasculature, and the associations included several cardiometabolic disease variants and risk factors. Further research is needed to investigate the potential of the retinal vessel tortuosity as a clinically relevant biomarker for cardiovascular disease and metabolic syndrome.
- Enriched pathways include a well-known therapeutic target for ocular diseases (VEGFA-VEGFR2) affecting tissue remodeling. We highlight several transcription factors as interesting targets for further experimentation.

## INTRODUCTION

Cardiovascular diseases (CVD) are the leading cause of death in developed countries[1–3] and a major societal health burden. Though several risk factors for CVD development, such as age, smoking, and hypertension, have been firmly established, the degree of importance of vascular properties as risk factors is unclear. Retinal fundus photos allow non-invasive in-vivo assessment of the vascular system of the superficial inner retina, i.e. the central and branch veins and arteries plus the venules and arterioles. These vessels are composed of tightly sealed endothelial cells (ECs), forming the inner blood-retina barrier (BRB), encased by smooth muscle cells (SMCs) form to the vessel wall [4,5]. Automatic segmentation of retinal vessels in fundus images is well established, and is commonly used in standard clinical care to screen and diagnose ocular and systemic diseases [6]. In diabetes, for example, hyperglycemia induces damage to the ECs and pericytes of the inner BRB contributing to retinal edema and hemorrhage [7].

Pathological changes in the retinal vessels often coincide with those in the microvasculature of other organs and may precede the progression of systemic vascular diseases. The retinal vasculature can provide insights into neuro-degenerative diseases, such as Alzheimer’s, Parkinson’s, and vascular dementia [8–12]. In addition, abnormalities in retinal parameters are of diagnostic value for systemic diseases, including increased risk of diabetes [13–15], obesity [16] and CVD [17,18] (such as stroke [19–22], coronary heart disease [23], peripheral artery disease [24], hypertension [21,25–33], atherosclerosis [19,21,34], myocardial infarction [35,36], and nephropathies [37,38]).

In recent years, genome-wide association studies (GWAS) have been used to link genes with phenotypes extracted from fundus images, such as vessel size [39,40], optic disc morphology [41,42], vascular density [43], fractal dimensions [43] and vessel tortuosity [44]. The diameter of the retinal microvasculature was associated with genes TEAD1, TSPAN10, GNB3 and OCA2 [39]. A recently published study [43] on vascular density and fractal dimensions reported 7 and 13 single nucleotide polymorphism (SNPs) associated with these traits respectively, including OCA2, MEF2C and GNB3. Retinal vessel tortuosity has been associated with SNPs that map to the genes ACTN4 and COL4A2 [44] Tortuosity of the vasculature was reported in the context of CAD [45] and connective tissue disease [46]. These results demonstrated that GWAS can reveal genes with a potential role in modulating vascular properties and related pathomechanisms, though their power has been often limited.

Here, we report the results of the largest GWAS on vessel tortuosity to date. Our study was motivated by the clinical relevance of this trait to diseases [9,13,28,46–48], as significant associations were already reported in smaller sample sizes, making further discoveries likely. We used images and genotypes from 62 751 subjects in the UK Biobank and from 397 and 512 subjects of the much smaller, yet independent, population-based cohorts SKIPOGH [49,50] and *OphtalmoLaus* [51]. We constructed an automated image analysis pipeline to extract retinal tortuosity from these data as a biomarker. We report the correlation with patient records, SNPs, genes, pathways, tissue expression, pathomechanisms, and causal effects associated with this biomarker.

## METHODS

### Data: genotypes, phenotypes and fundus images

The UK Biobank is a population-based cohort of approximately 488 000 subjects with rich, longitudinal phenotypic data and a median 10-year follow-up [52,53]. We analyzed 173 837 retinal fundus images from 84 825 individuals. Genotyping was performed on Axiom arrays for a total of 805 426 markers, from which approximately 96 million genotypes were imputed. We used the subset of 15 599 830 SNPs that had been assigned an rsID. We performed an additional quality control step by filtering out SNPs with MAF < 5×10^−4^ (which translates to an expected minimum number of ∼60 individuals expected to have at least one minor allele). Finally, we applied a filtering procedure [54] to remove SNPs with imputation quality < 0.3. In addition to genomic information, we obtained phenotypic information from the patient records: type-2 diabetes, angina, myocardial infarction, deep vein thrombosis (DVT), stroke, hypertension and smoking status. Age, sex, and principal components of genotypes were used to correct for biases in the genetic associations.

We performed replication via a meta-analysis of two independent, population-based cohorts: the *Swiss Kidney Project on Genes in Hypertension* (SKIPOGH) [49,50] and *OphtalmoLaus* [51]. SKIPOGH is a family-based, cross-sectional study exploring the role of genes and kidney hemodynamics in blood pressure regulation and kidney function in the general population, comprising 1 054 genotyped individuals. 1 352 retinal fundus images were available from 518 participants. The genotyping was performed with the Illumina Omni 2.5 chip. *OphtalmoLaus* is a sub-study of *Cohorte Lausannoise* (*CoLaus*), a population-based cohort comprising 6 188 genotyped individuals. 7 252 fundus images were available from 1 015 subjects. *CoLaus* has as its objective to investigate the epidemiology and genetic determinants of CVD risk factors and metabolic syndrome: participants were phenotyped accordingly. The genotyping was performed using the 500K Affymetrix chip technology.

### Automated analysis of color fundus images and quality control

We extended the software ARIA [55] to perform batch segmentation and positional annotation of blood vessels, using the default parameters [56]. We applied a quality filter based on the total length of the vasculature and on the number of vessels (see Supplemental Text 1). Roughly two out of three images passed this strict quality control (116 639 out of 173 837 in the UK Biobank). Based on ARIA’s vessel annotations, we calculated a tortuosity measure known as the *distance factor* (DF) [57], defined as:

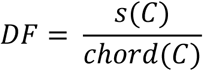

where the total vessel length, *s*(*C*), is divided by the Euclidean distance between the vessel segment endpoints, *chord*(*C*). DF is referred to in a recent review as the arc over chord ratio [58]. In addition to DF, we also calculated six other tortuosity phenotypes based on alternative measures using integrals over the curvature along the vessel (see Supplemental Text 2).

We phenotyped each individual by calculating median retinal tortuosities, then averaging the values derived from the left and right eye, when available (for the resulting distribution, refer to Supplemental Text 3).

### Deep Learning classification of arteries and veins

We calculated pixel-wise artery and vein classifications using the Deep Learning algorithm *Little W-Net* [59]. For each vessel segment recognized by ARIA, we used the difference between pixels classified as arterial and venous as a score that was required to be positive or negative for the segment to be annotated as artery or vein, respectively. On a set of 44 images, manually annotated by an ophthalmologist, we obtained an area under the curve of 0.93 and an accuracy of 0.88. Thus, we performed vessel type classification for the entire set of retinal fundus images, computing artery- and vein-specific tortuosity values (see Supplemental Text 4).

### Genome-wide association analyses

We ran genetic association studies on tortuosity of arteries, of veins, and combining both vessel types. We used BGENIE [60], applying linear regression to confounder-corrected, quantile-quantile normalized, retinal vessel tortuosity on the genotypes of the matching subjects imputed to a panel of approximately 15M genetic variants. In order to account for confounding effects [61], the following variables were provided as covariates: age, sex, PC of the genotypes (we considered only PCs with a significant correlation to tortuosity, namely 1, 2, 5, 6, 7, 8, 16, 17 and 18). We applied a Bonferroni threshold of 5×10^−8^. A list of independent SNP was obtained by performing linkage disequilibrium (LD) pruning using the LDpair function of the R package LDlinkR [62]. Two SNPs were considered independent if they had LD *r*^*2*^<0.1 or were more than 500K bases apart (see <u>Supplemental Dataset 1</u>).

### Replication meta-cohort

As the SKIPOGH cohort includes subjects with a high degree of relatedness, we used the EMMAX function of the EPACTS software [63] and the kinship matrix in the model to account for family structure. We also included the recruitment center as a covariable. For the GWAS on the *OphtalmoLaus* cohort, we used the same parameters and tools as for the discovery cohort. Results from SKIPOGH and *OphtalmoLaus* were meta-analyzed using an inverse-variance weighting scheme for the respective effect sizes as well as a random-effects model (Supplemental Text 5).

To keep the multiple hypotheses testing burden low, we took a candidate gene approach. Specifically, we used the Benjamini–Hochberg procedure [64], which applies no multiple hypothesis testing correction for the top hits, and then corrects the association of the candidate locus of rank *n* for *n* tests.

### Heritability estimates

We used LD Score Regression [65] to estimate the SNP-based heritability of our tortuosity measures.

### Novel method for gene-based tests

We used *PascalX* [66], a novel high-precision pathway scoring algorithm that we developed building on our *Pascal* [67] tool, to aggregate SNP-wise summary statistics into gene scores using a sum of □^2^ statistics: *PascalX* takes into account LD by effectively transforming the sum of □^2^ from all SNPs within the gene window into a new basis of independent “Eigen-SNPs” corresponding to a weighted sum of □^2^ statistics. Using multiple-precision arithmetics, *PascalX* computes the corresponding null cumulative probability distribution to essentially arbitrary precision, while other tools usually only approximate the underlying distribution. We thus computed p-values up to a precision of 10^−100^, allowing for accurate scoring of genes with contributions from extremely significant SNPs, which become increasingly frequent in highly powered GWASs such as this one.

We used the following configurations: We computed gene scores from SNPs lying within a window of 50kb before the transcription start site and 50kb after the transcript end. The annotation of the gene positions was based on the Genome Reference Consortium Human genome build 37 (GRCh37/hg19) downloaded from the Ensembl biomart [68]; we considered only protein-coding and lincRNA genes. The reference panel from the UK10K project [69] was used to estimate the SNP-SNP correlations (LD effects). *PascalX* uncovered 265 significant genes (after Bonferroni correction for 25 489 gene-based tests *p* < 0.05 / 25489 ≃2.0×10^−6^).

### Gene set enrichment

We used *PascalX* [66] to compute gene set enrichment scores based on ranking derived from the gene-based tests. As a large number of genes have inflated p-values in highly powered GWASs, this ranking approach was more conservative. We first computed scores for 2 868 canonical pathways (BioCarta, KEGG, PID, Reactome, and WikiPathways), then extended our analysis to the 31 120 pathways in MSigDB (v7.2) [70]. To adjust for statistical dependence and co-expression, genes that are less than 100kb apart were “fused” (i.e. considered as single entities termed “fusion genes” [67]).

### Tissue-wide gene expression analysis

We performed tissue-wide gene expression analysis using *PascalX* [66] on the whole GTEx [71] (v8) dataset, comprising 54 tissues. We defined gene sets based on the significant genes from each of our three GWAS on DF tortuosity (artery, vein and combined). *PascalX* was used to perform an enrichment analysis that indicated whether these sets were over-expressed in any particular tissue. *PascalX* corrected for the co-expression of gene sub-clusters within each gene set by merging nearby genes to fusion genes. We computed the fusion genes expression values in transcripts per kilobase million from the raw read counts. These values values were made uniform via ranking, transformed to □^2^-distributed random variables, summed, and tested against a □^2^ distribution with as many degrees of freedom as there were “fusion genes” in each set. We applied a Bonferroni threshold: *p* = 0.05 / 54 = 9.2×10^−4^.

### Shared genetic architecture with disease

We computed the overlap between DF tortuosity SNPs (from the combined-vessel GWAS) and disease-related SNPs. To this end, we first identified which of the independent SNPs in the combined-vessel GWAS were listed in the GWAS Catalog [72]. We then extended this analysis by considering DF tortuosity SNPs in LD (*r*^*2*^ > 0.8) with disease-related SNPs in the GWAS Catalog.

### Mendelian randomisation analysis

We performed two-sample bidirectional Mendelian randomisation [73,74] to search for evidence of causal effects between DF tortuosity (from the combined-vessel GWAS) and the following traits: body mass index (BMI), coronary artery disease (CAD), systolic blood pressure (SBP), and lipid traits, namely high-density lipoprotein, low-density lipoprotein (LDL), total cholesterol, and triglycerides. For each trait, we used independent (*r*^*2*^ < 0.01) significant (*P* < 5×10^−8^) SNPs as instrumental variables. All summary statistics (estimated univariate effect size and standard error) originated from the most recent meta-analyses (not including UK Biobank individuals) and were downloaded from the publicly available NIH Genome-wide Repository of Associations Between SNPs and Phenotypes [75]. We only used SNPs on autosomal chromosomes available in the UK10K reference panel [69], which allowed us to estimate the LD among these SNPs and prune them. We removed strand ambiguous SNPs. Causal estimates were based on the inverse variance weighted method [76] and calculated using the Mendelian randomisation R package [77].

### Code Availability

Code for all computations is available on request to the corresponding author.

## RESULTS

### Baseline characteristics and tortuosity quantification

Following quality control measures, we analyzed 116 639 images from 62 751 subjects of the UK Biobank (mean±SD age = 56±8 years; 35 098 females at birth [54%]; 4 618 smokers [7%]). We analyzed 1 352 images from 379 subjects of the SKIPOGH cohort (mean±SD age = 48±16 years; 211 females [53%]; 107 smokers [27%]). We analyzed 7 254 images from 512 subjects of the *OphtalmoLaus* cohort (mean±SD age = 51±10 years; 270 females [53%]). Baseline characteristics and disease prevalence are presented in Supplemental Text 6. For an overview of our pipeline see Figure 1.

**Figure 1.**
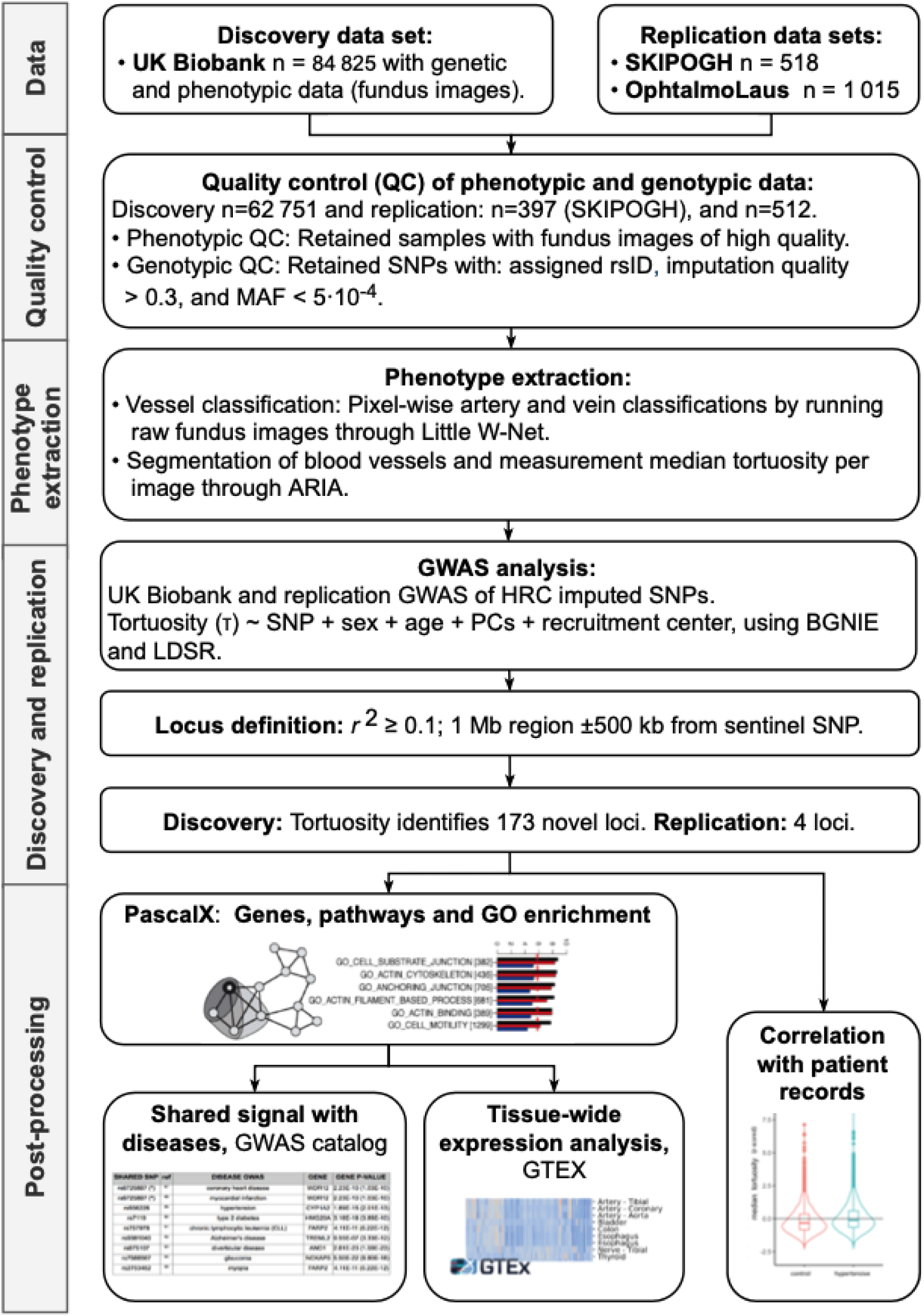
Pipeline and results. Relevant phenotypes, genotypes and fundus images were collected from the UK Biobank, *OphtalmoLaus* and SKIPOGH. After quality control, the images were processed by deep learning, classifying arteries and veins. A range of tortuosity measures were then calculated, which provided the phenotypes for the GWASs. The primary results were 173 novel genetic trait loci. The association signals pointed to a shared genetic architecture of retinal tortuosity and disease (metabolic syndrome and CVD). Their aggregation on annotated gene-sets identified relevant pathways and GO terms. Tissue-wide expression analysis revealed expression in the arteries and heart. Correlation analysis revealed associations between retinal tortuosity and cardiometabolic diseases.

The distributions of DF tortuosity were similar across cohorts: long-tailed, left-skewed, with means ranging from 1.030 (UK Biobank) to 1.034 (*OphtalmoLaus*). DF was higher in the elderly population (Cohen’s d = 0.49, *p =* 1×10^−195^), while sex had limited impact (Cohen’s d = 0.049, *p =* 9×10^−10^). Overall, DF was higher in veins (Cohen’s d = 0.13, *p =* 9×10^−142^). For details about the stratified analysis of the DF phenotype in the UK Biobank see Supplemental Text 3.

We extracted six additional tortuosity measures based on alternative mathematical definitions. Correlations analysis and dimensionality reduction in terms of principle components showed that the DF is most similar to the path integral of the squared curvature (*τ*_3_) and least similar to the path integral of the curvature (*τ*_2_). The other alternative measures (*τ*_4-7_) were similar to each other, very different from *τ*_2_ and of intermediate similarity to the DF and *τ*_3_ (see Supplemental Text 2).

### Vessel tortuosity correlates with disease status

We found DF tortuosity (from the combined-vessel GWAS) to be associated with angina, myocardial infarction, stroke, DVT and hypertension. Analyzing arteries and vein separately, we found that the DF of arteries was significantly associated with hypertension (*beta* = 0.19, *p =* 3×10^−56^) and angina (*beta* = 0.09, *p =* 6×10^−4^), but not with myocardial infarction, stroke or DVT. In the case of veins, the DF was significantly associated with hypertension (*beta* = 0.25, *p =* 7×10^−99^), angina (*beta* = 0.18, *p* = 2×10^−10^), myocardial infarction (*beta* = 0.12, *p =* 2×10^−4^), stroke (*beta* = 0.16, *p =* 5×10^−5^) and DVT (*beta* = 0.11, *p =* 5×10^−4^). For predictive power over disease status, see Supplemental Text 7.

### Vessel tortuosity GWASs identify 173 novel loci

We identified 7 072 significantly associated SNPs in the combined-vessel GWAS on DF tortuosity in the UK Biobank (<u>Supplemental Dataset 4A</u>). The vessel type specific GWAS resulted in 6 563 significantly associated SNPs for arteries, and 2 896 SNPs for veins (<u>Supplemental Dataset 4B</u> and <u>4C</u>). We applied LD pruning, identifying 128 independent loci in the combined-vessel GWAS, 116 in the artery-specific GWAS, and 63 in the vein-specific GWAS. Accounting for overlap between these sets (see Supplemental Text 9), we obtained a total of 175 independent lead SNPs (see Figure 2a-c). The top 10 SNPs are listed in Table 1, ordered by significance (for complete listings, see Supplemental File 1). Among the significantly associated variants, rs1808382 and rs7991229 had been previously reported [45] (Supplemental Text 8), whereas the remaining 173 independent lead SNPs represented novel loci associated (see Supplemental File 5).

**Table 1.** Top retinal tortuosity SNPs. The 10 most significant DF tortuosity SNPs, ordered by p-value. For full results, refer to the list of 175 independent lead SNPs in Supplemental Dataset 1. **Chr**: chromosome; **SNP**: rsIDs of the single nucleotide polymorphism; **EA**: effect allele; **RA**: reference allele; **freq**: allele frequency of effect allele; **beta**: effect size estimate; **-log**_**10**_ ***p***: normalized p-value in the discovery cohort; **GWAS type**: vessel type to which the signal applied.

| Chr | SNP | EA | RA | freq | beta | -log<sub>10</sub> p | GWAS type |
| --- | --- | --- | --- | --- | --- | --- | --- |
| 13 | rs9559797 | G | C | 0.580 | -0.162 | 182.4 | artery and vein |
| 19 | rs16972767 | G | A | 0.473 | -0.150 | 164.6 | artery and vein |
| 4 | rs17008193 | T | C | 0.403 | -0.089 | 55.0 | artery and vein |
| 7 | rs187691758 | A | G | 0.005 | 0.627 | 53.2 | vein |
| 4 | rs12506823 | G | A | 0.406 | 0.083 | 47.9 | artery and vein |
| 2 | rs2571461 | T | G | 0.601 | -0.082 | 47.1 | artery and vein |
| 15 | rs12913832 | A | G | 0.744 | 0.080 | 43.6 | artery and vein |
| 12 | rs11045245 | A | G | 0.375 | 0.073 | 37.3 | artery |
| 5 | rs784420 | A | G | 0.281 | 0.078 | 37.0 | artery and vein |
| 4 | rs11727963 | G | A | 0.166 | 0.092 | 35.1 | artery |

**Figure 2.**
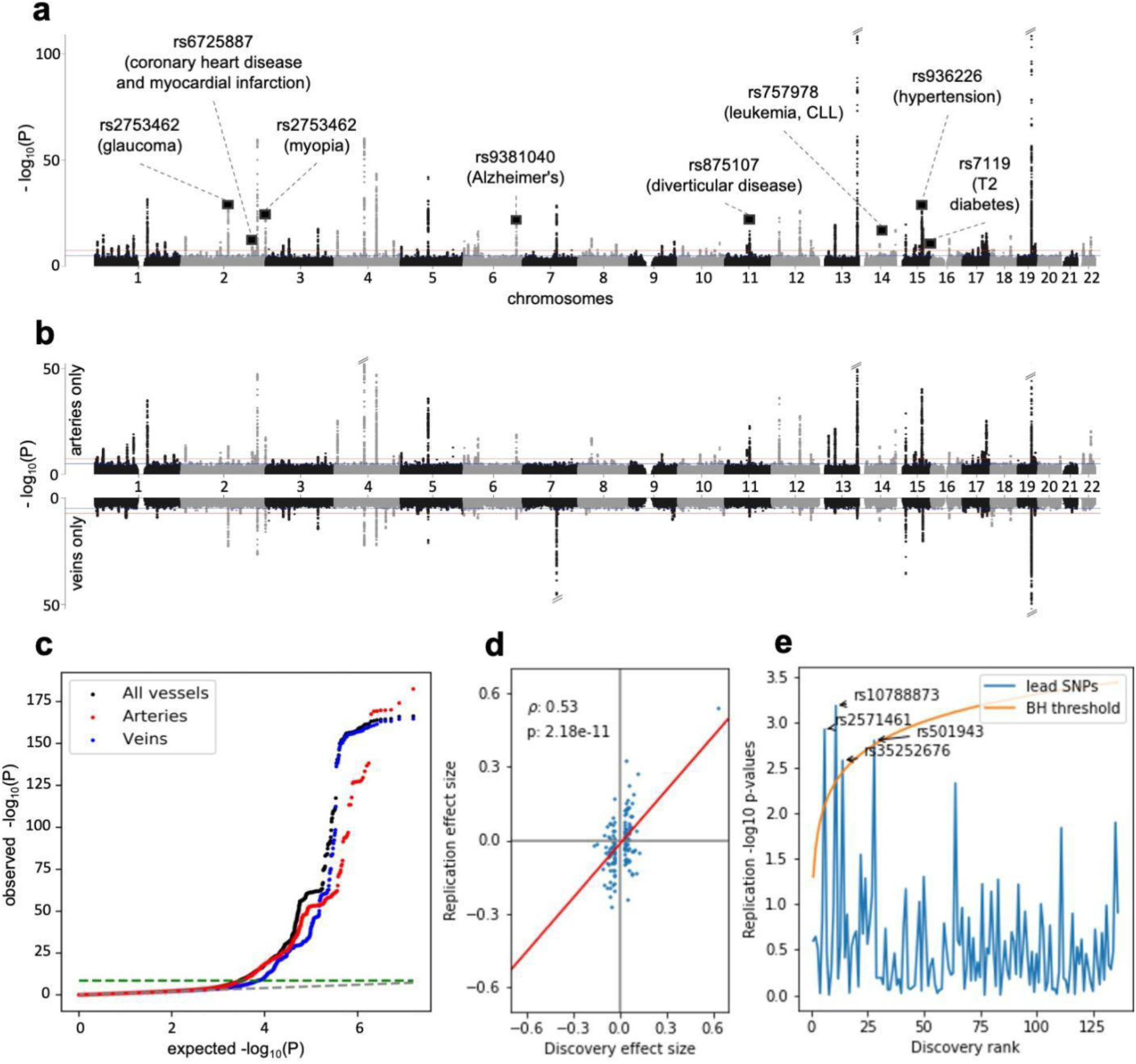
SNP p-values and effects. **a**, Manhattan plot of Genome-Wide Association Study (GWAS) of retinal vessel tortuosity, combining all vessel types (both arteries and veins). The red line indicates the genome-wide significance level after Bonferroni correction (*p* = 5×10^−8^). Oblique dashes on top of peaks mark extremely significant p-values that have been cropped. Squares mark the position of disease SNPs (see Table 4). The trait was corrected for phenotypic variables which showed a statistically significant association, i.e.: age, sex, and a subset of principal components of genotypes. **b**, Manhattan plots of the vessels-specific GWAS (artery-specific on top, vein-specific at the bottom). Confounder correction, significance level and cropping of extremely significant p-values as in the (a). **c**, GWAS q-q plot: arteries in red, veins in blue, combined-vessels signal in black; the genome-wide significance level is represented as a green dashed line. **d**, Statistically significant correlation between the measured effect sizes in the discovery cohort (UK Biobank, n = 62 751) and replication meta-cohort (SKIPOGH plus *OphtalmoLaus*, n = 911). We considered all lead (independent) SNPs in the UK Biobank. We could find 136 with matching rsIDs in the replication meta-cohort, 90 of which had the same sign of their effect size estimate in the UK Biobank. The resulting Pearson correlation is *r =* 0.53; *t=*7.28, *p =* 1.18×10^−11^. **e**, Benjamini-Hochberg procedure on discovery lead SNPs from the UK Biobank yields 4 hits in the replication cohort.

### Heritability of DF is larger than for other tortuosity measures

The SNP-based heritability differed substantially across tortuosity measures, with DF receiving the highest estimate (h^2^_SNP_= 0.25, SE = 0.025). This was approximately twice the heritability estimate of the six alternative curvature-based measures (0.11 ≤ h^2^_SNP_≤ 0.13, 0.011 ≤ SE ≤ 0.012, Supplemental Text 2). We did not observe any significant genomic inflation (see Table 2). Heritability also varied depending on vessel type (h^2^ _SNP_ = 0.23 [SE = 0.020] for arteries, and h^2^ _SNP_ = 0.15 [SE = 0.021] for veins). The distribution of the DF phenotype for each vessel type is shown in Supplemental Text 3.

**Table 2.** SNP-based heritability. h^2^_SNP_: portion of phenotypic variance cumulatively explained by the SNPs; **lambda GC**: inflation, measure of the effect of confounding and polygenicity acting on the trait; **intercept**: LD score regression intercept (values close to 1 indicates little influence of confounders, mostly of population stratification); **ratio**: ratio of the proportion of the inflation in the **mean Chi**^**2**^ that is not due to polygenicity (a ratio close to, or smaller than, 0 is desirable as it indicates low inflation from population stratification). SE are given in parentheses.

| GWAS type | h<sup>2</sup>SNP | lambda GC | mean Chi<sup>2</sup> | intercept | ratio |
| --- | --- | --- | --- | --- | --- |
| combined-vessel | 0.25 (0.025) | 1.14 | 1.31 | 1.01 (0.01) | 0.03 (0.03) |
| artery | 0.23 (0.020) | 1.12 | 1.27 | 1.00 (0.01) | < 0 |
| vein | 0.15 (0.021) | 1.10 | 1.18 | 1.00 (0.01) | < 0 |

**Table 3.** Top retinal tortuosity genes. The 15 most significant DF tortuosity genes, for each GWAS (combining all vessels, considering only arteries, and only veins). P-values were computed by *PascalX [66]* (precision cutoff: 1×10^−100^). For full results, refer to Supplemental Dataset 6A / 6B / 6C.

| Gene | Chr | base pair | -log<sub>10</sub> p combined | -log<sub>10</sub> p artery | -log<sub>10</sub> p vein |
| --- | --- | --- | --- | --- | --- |
| ACTN4 | 19 | 39,138,289 | > 100 | > 100 | > 100 |
| CAPN12 | 19 | 39,220,827 | > 100 | > 100 | > 100 |
| EIF3K | 19 | 39,109,735 | > 100 | > 100 | > 100 |
| LGALS7 | 19 | 39,261,611 | > 100 | > 100 | > 100 |
| LGALS7B | 19 | 39,279,851 | > 100 | > 100 | > 100 |
| COL4A2 | 13 | 110,958,159 | > 100 | > 100 | 9.5 |
| LGALS4 | 19 | 39,292,311 | > 100 | 34.5 | > 100 |
| MAP4K1 | 19 | 39,078,281 | > 100 | 34.3 | > 100 |
| TNS1 | 2 | 218,664,512 | > 100 | 32.7 | 16.9 |
| ECH1 | 19 | 39,306,062 | > 100 | 15.3 | > 100 |
| AC104534.3 | 19 | 39,310,806 | > 100 | 14.0 | > 100 |

**Table 4.** Pleiotropic disease-variants. List of variants identified in the tortuosity GWAS (combined-vessel analysis) which were found to be associated with a disease outcome or risk factor in an independent study. We report only exact variants (same rsID in both tortuosity and disease GWAS), which we could confidently map to a gene. Gene p-values were computed by *PascalX* [66]. Variants associated with more than one disease are marked by a star (*)

| Shared SNP | Disease GWAS | Gene | $-\log_{10} p$ | ref |
| --- | --- | --- | --- | --- |
| rs875107 | diverticular disease | ANO1 | 22.5 | [105] |
| rs7588567 | glaucoma | NCKAP5 | 21.2 | [106] |
| rs7119 | type 2 diabetes | HMG20A | 17.5 | [107] |
| rs936226 | hypertension | CYP1A2 | 14.7 | [108] |
| rs757978 | chronic lymphocytic leukemia (CLL) | FARP2 | 10.4 | [109] |
| rs2753462 | myopia | FARP2 | 10.4 | [110] |
| rs6725887 (*) | coronary heart disease | WDR12 | 9.6 | [111] |
| rs6725887 (*) | myocardial infarction | WDR12 | 9.6 | [112] |
| rs9381040 | Alzheimer's disease | TREML2 | 6.0 | [113] |
| rs11083475 | heart rate (rhythm disorders) | ACTN4 | > 100 | [114] |
| rs9555695 | waist-hip ratio (obesity) | COL4A2 | > 100 | [115] |
| rs2571445 | lung function (pulmonary disease) | TNS1 | > 100 | [116] |
| rs3791979 | intraocular pressure (open angle glaucoma) | TNS1 | > 100 | [117] |
| rs17263971 | eGFR (Chronic Kidney Disease) and retinal dysfunction | SYNPO2 | 28.7 | [79,118] |
| rs35252676 | pulse pressure (CVD) | LHFPL2 | 26.6 | [119] |
| rs1378942 (*) | diastolic blood pressure (CVD) | CSK | 23.4 | [120] |
| rs1378942 (*) | mean arterial pressure (CVD) | CSK | 23.4 | [121] |
| rs17355629 | pulse pressure (CVD) | LRCH1 | 19.6 | [122] |
| rs7655064 | waist-hip ratio (obesity) | MYOZ2 | 14.5 | [115] |
| rs6495122 | diastolic blood pressure (CVD) | CPLX3 | 14.5 | [123] |
| rs12913832 | intraocular pressure (open angle glaucoma) | HERC2 | 12.3 | [124] |
| rs9303401 | cognitive ability (mental disorders) | PPM1E | 10.01 | [125] |

### Replication of lead SNPs and genes in a small meta-cohort

We replicated four of the lead SNPs from the UK Biobank GWAS (see Figure 2e), and discovered three associated genes (TNS1, AC011243.1, LHFPL2; see Figure 3e) [64]. Given the limited sample size of the replication meta-cohort (n *=* 911), we performed additional analyses on trend of effect sizes. We observed a Pearson correlation of *r* = 0.53 (*p* = 1.2×10^−11^) between the effect size estimates in the two studies (see Figure 2d, Figure 3d and Supplemental Text 5).

**Figure 3.**
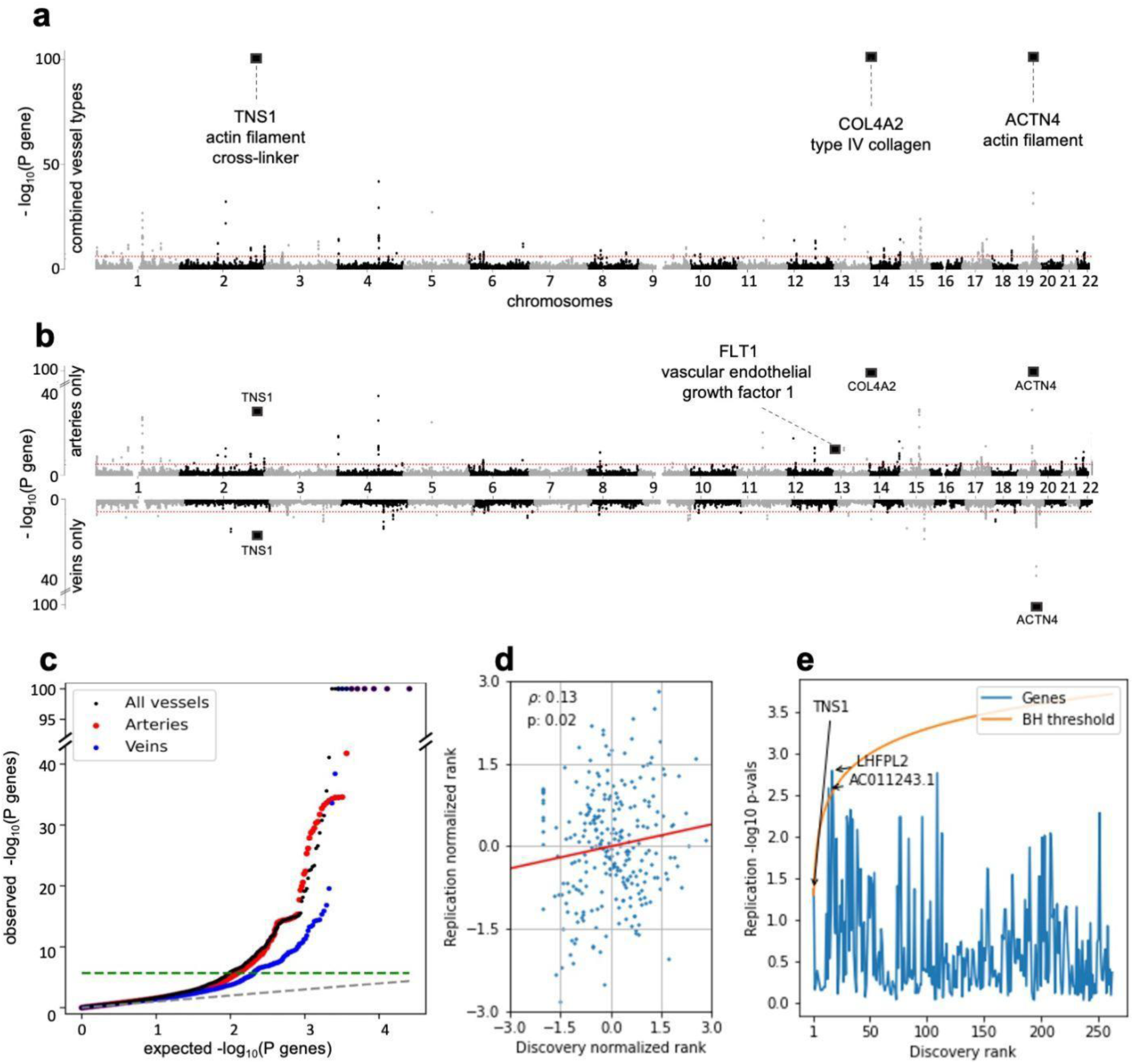
Gene p-values and replication scores. **a**, Gene-based Manhattan plot of retinal vessel tortuosity, combining all vessel types (both arteries and veins). Gene-based tests were computed by *PascalX* [66]. The red line indicates the genome-wide significance level after Bonferroni correction (*p =* 5×10^−8^). Squares mark the position of particularly relevant genes (see corresponding Results section). **b**, Gene-based Manhattan plots of the vessels-specific GWAS (artery-specific on top, vein-specific at the bottom). **c**, q-q plot of gene p-values: arteries in red, veins in blue, combined-vessel signal in black; the genome-wide significance level is represented as a green dashed line. **d**, Statistically significant correlation between q-q normalized genes’ p-values in the discovery (UK Biobank) and in the replication meta-cohort (SKIPOGH + *OphtalmoLaus*). Only genes that were significant in the discovery cohort were considered. The resulting Pearson correlation is *r =* 0.13; *p =* 0.02. **e**, Benjamini-Hochberg generates 3 hits in the replication meta-cohort. Only genes that were significant in the discovery cohort were considered.

### Tortuosity genes and pathways affect vascular tissue remodeling and angiogenesis

Mapping the SNP-wise association signals onto genes (Methods) we identified 265 significant genes in the discovery GWAS combining vessel types, 203 in the artery-specific GWAS, and 123 in the vein-specific GWAS. Accounting for overlap between these sets (see Supplemental Text 9), we obtained a total of 312 genes (see Figure 3a-c). Top genes are reported in Table 3 (for a complete listing, see <u>Supplemental Dataset 6A / 6B / 6C</u>). A large fraction of these genes carried annotations related to vessel integrity, vascular tissue remodeling and angiogenesis. Specifically, we identified a cluster of highly significant genes on chromosome 19, including *ACTN4* (related to actin filament bundling), *TNS1* (cross-linking of actin filaments), and *CAPN12* (involved in structural integrity to blood vessel walls). This locus also included three genes involved in adhesion to the connective tissue [78]: *LGALS7, LGALS7B* and *LGALS4*. We also replicated the highly significant association of tortuosity with two type IV collagen genes, *COL4A2 and COL4A1* [44]. *SYNPO2* (related to actin polymerisation, vascular injury [79] and ocular growth [80], also received a highly significant association. Finally, among the artery-specific genes, we found *FLT1* coding for VEGFR1, which plays a role in vessel formation and vascular biology [81]. (See Discussion for further details and interpretation of these results.)

Gene set enrichment (Methods) yielded 78 significant sets in total (see Figure 4), with the strongest signals arising from the combined and artery-specific analysis (see Supplemental Text 9 and <u>Supplemental Dataset 7A / 7B / 7C</u>). Similarly to genes, many of the pathways pointed to specific biological processes, cellular components, and molecular functions related to vessel integrity and remodeling. These included “human retinal fibroblasts”, “vascular smooth muscle cells” (both in the kidney and the neuroepithelium), and “epithelium development”. We also observed a pathway related to “vascular endothelial growth factors”, VEGFA-VEGFR2, which is a well-known therapeutic target for ocular diseases. We highlight several transcription factors and binding motifs for further experimentation (see Figure 4b). The role of integrity and development of blood vessels for tortuosity was supported by the enrichment of several GO terms such as “circulatory system development”, “anatomical structure morphogenesis” and “tube development”. The enriched terms “cell-substrate junction”, “anchoring junction”, “actin” and “actomyosin” revealed some of the molecular players involved. (See Discussion for more details).

**Figure 4.**
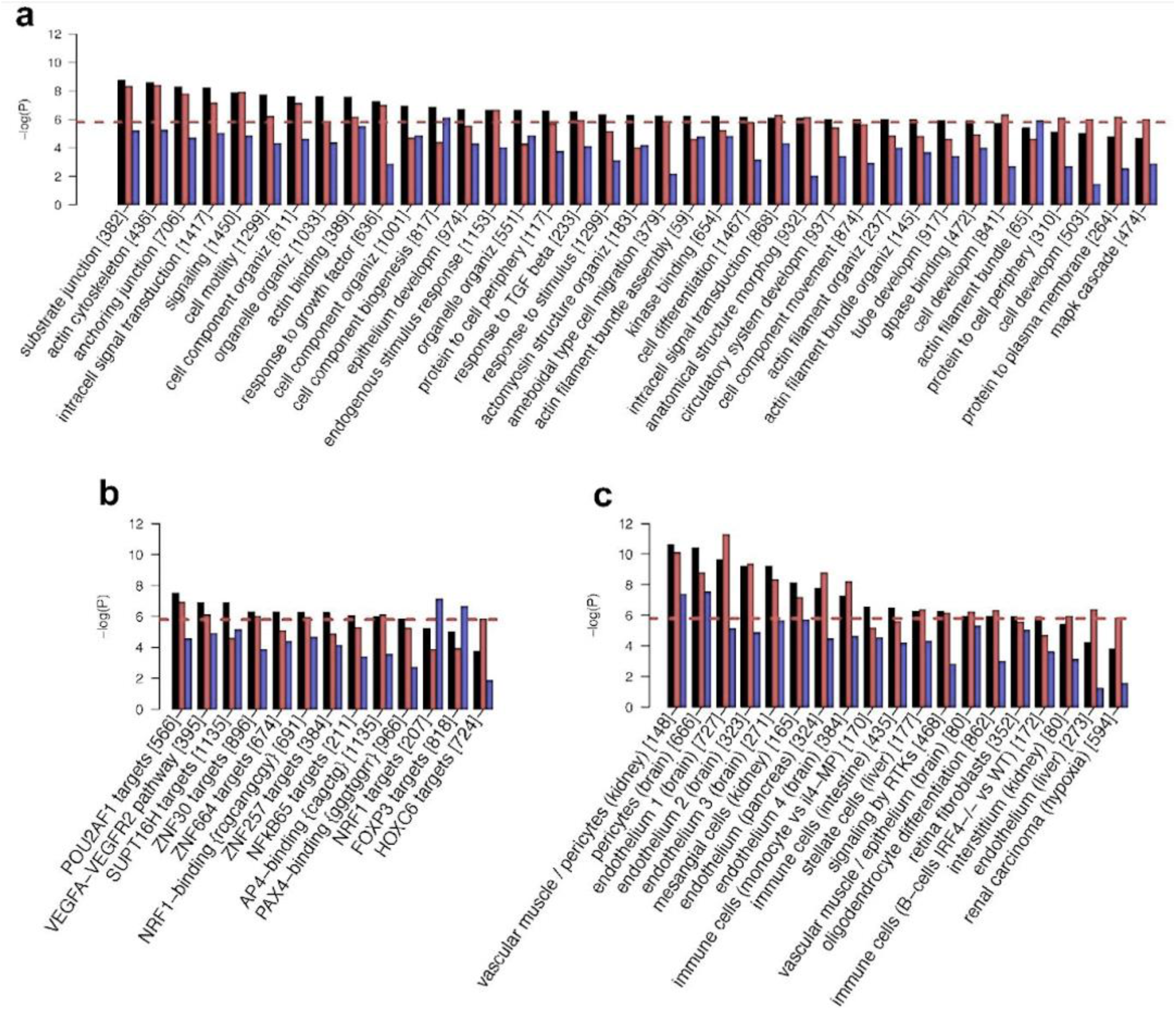
Enriched pathways and gene-sets. Arteries in red, veins in blue, combined-vessel signal in black: scores for 31 120 gene-sets in MSigDB (v7.2) [103] were calculated by *PascalX* [66]. Only gene-sets for which significance was reached by at least one GWAS are shown. The red dashed line indicates Bonferroni-threshold (-log_10_ *p =* 5.7). The number of genes in each set is indicated in squared brackets. Gene-set names have been shortened and some redundant GO categories are not shown. For details, refer to the extended plot in Supplemental Text 13. **a**, Enrichment in GO categories. **b**, Enrichment in pathways referring to a particular molecule (typically a transcription factor) and/or binding motif. **c**, Enrichment in gene-set obtained from transcriptomic analysis of tissues of treated cell types.

Compared to the DF analysis, the alternative tortuosity measures had lower heritability and fewer enriched genes and pathways. However, some were unique and disease-relevant, such as a pathway related to “abnormal cardiac ventricle morphology” (see Supplemental Text 2).

### Tortuosity genes are overexpressed in arteries and heart tissues

Performing enrichment analyses across expression data from 54 tissues, we found that tortuosity genes were overexpressed in three types of arteries (i.e., aorta, tibial artery and coronary artery), two heart tissues (i.e. ventricle and atrial appendage), and, less significantly, fibroblasts and muscular tissues. The profile of enrichment significance values across tissues for tortuosity genes detected by combined-vessel type GWAS analysis is more similar to that of the artery-specific GWAS than that of vein-specific one (see Figure 5), which did not result in any significant tissue associations (for a strict Bonferroni threshold of *p* = 0.05/54 = 9.2×10^−4^).

**Figure 5.**
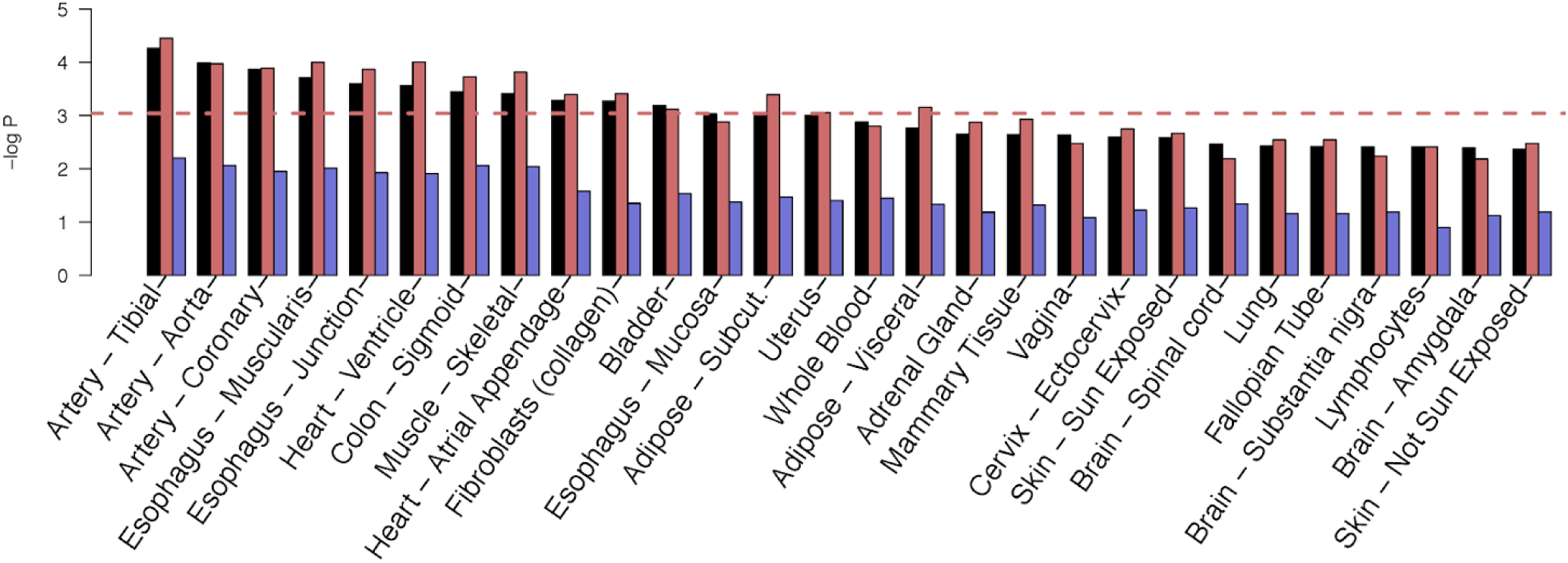
Tissue expression results. Arteries in red, veins in blue, combined-vessel signal in black: tissue-specific gene expression analysis of GTEx (v8) [71] performed using *PascalX* [66]. We defined sets based on the significant genes from each of the three GWAS we carried out and asked whether they were over-expressed in a particular tissue.

### Tortuosity loci are known disease variants

Nine of the discovered tortuosity loci had been previously reported as disease variants that mapped to specific genes (Table 4): four loci were linked to vascular diseases (coronary heart disease, myocardial infarction, arterial hypertension, and diverticular disease), two loci were linked to ocular diseases (glaucoma and myopia), and three loci were linked to other systemic diseases (chronic lymphocytic leukemia, type 2 diabetes, and Alzheimer’s disease). Similarly, we identified 12 loci influencing both tortuosity and disease risk factors. We also uncovered 26 additional disease variants that have not been confidently mapped to a specific gene (see Supplemental Text 10).

### Genetic overlap with cardiometabolic risk factors

We expanded our analysis of disease variants to SNPs belonging to the same LD block (Figure 6). We observe a sizable number of tortuosity-associated variants that overlap with CVD (54 SNPs). Several traits related to metabolic syndrome also stand out: blood pressure (55 SNPs for SBP, 49 for DBP, 15 for pulse pressure), blood cholesterol levels (54 SNPs), BMI (54 SNPs), blood pressure linked to alcohol intake and smoking (44 SNPs for SBP + alcohol, 27 for DBP + alcohol) and type2 diabetes (5 SNPs). In addition, other CVD risk factors share a high number of variants associated with tortuosity, such as protein levels (27 SNPs) and type1 diabetes (9 SNPs). Finally, we detected an overlap with various eye morphology traits, including optic disc morphometry (40 SNPs).

**Figure 6.**
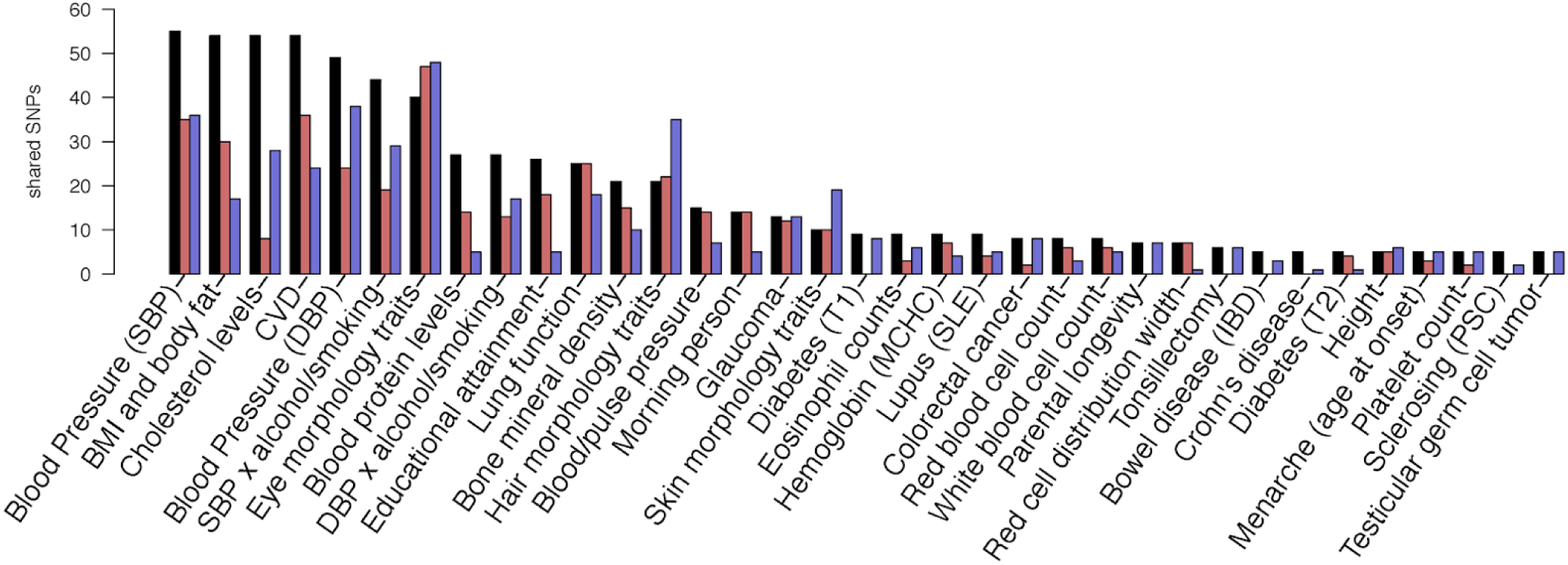
Overlap in genetic signals with diseases and other complex traits. Arteries in red, veins in blue, combined-vessel signal in black: number of variants shared with other traits reported in the GWAS Catalog [72] (also considering SNPs in high LD with the lead SNP, *r*^*2*^ > 0.8). Only traits with at least 5 shared associations are included (for a full list, including rsIDs, refer to the <u>Supplemental Dataset 3</u>). The traits with the highest number of shared SNPs belong to metabolic syndrome (blood pressure, BMI, blood cholesterol levels) and CVD. This analysis was generated using FUMA [104].

### Causal effects between tortuosity, BMI and LDL

Using inverse-variance weighting MR, we observed that exposure to elevated (standardized) levels of LDL reduced the tortuosity of veins by 3% (*p =* 0.02) and arteries by 5% (*p =* 0.001). Conversely, increased venous (but not arterial nor combined) tortuosity reduced BMI by 4.4% (*p =* 0.01) (see Supplemental Text 11).

## DISCUSSION

Blood vessel tortuosity is a complex trait whose variation is induced in part during developmental angiogenesis and vascular differentiation and in part through vessel remodeling due to pathological processes in adult life. Both sources of variation are modulated by the environment, but also genetically through gene and regulatory variants that subtly modulate these processes. In order to better understand the involved genetic architecture we conducted the largest GWAS on retinal vessel tortuosity to date, identifying 173 novel loci and pinpointing genes and gene-sets enriched with these primary association signals. Leveraging the unprecedented number of hits, we performed MR that revealed the causal relationships between retinal tortuosity, BMI and blood lipids. This provides context for the considerable overlap we observed between variants associated with vessel tortuosity and cardiometabolic diseases as well as their risk factors. Our results were consistent with the overexpression of tortuosity genes in the aorta, tibial artery, coronary artery, and heart tissues. We found tortuosity genes to be involved in the development of blood vessels, the maintenance of vessel integrity and the remodeling as a consequence of disease processes.

### Vessel integrity

Several enriched GO categories that are integral to vessel development were enriched, namely “morphogenesis of anatomical structures”, “development of circulatory system”, and “tube development”. Similarly GO categories pertinent to the structural integrity of vessels and the stability of specific tissues were highlighted: “cell-substrate junction” and “anchoring junction” which are responsible for the mechanical attachment of a cell and its cytoskeleton to the extracellular matrix. Molecularly, “actin cytoskeleton”, “actin binding”, “actin filament bundle organization”, and “positive regulation of actin filament bundle assembly” highlighted the important role of actin.

Among the top hits, we found genes directly related to vessel integrity. The product of *ACTN4*, contributes to cell adhesion and to assembly of the tight junction by mediating actin filament bundling. The paralogues *COL4A1* and *COL4A2* provide structural support and elasticity to connective tissues by forming the hetero-trimer α1α1α2, which is the most abundant collagen in the basement membrane [82]. We found both *COL4A2* and *ACTN4* to be over-expressed in vascular tissues (see Supplemental Text 12). Two more genes with actin-related activity were also among our top hits: *TNS1*, which promotes cell migration and regulates angiogenesis [83], and *SYNPO2*, which is activated by actin polymerization, highly expressed in SMCs [79] and known to provide structural integrity to blood vessel walls [84]. Finally, we identified three genes coding for galectins, which are involved in adhesion to the connective tissue via modulation of cell-cell and cell-matrix interactions [78]: *LGALS7*, its paralog *LGALS7B* and *LGALS4*.

### Vessel remodeling

Pathological stresses such as inflammation, infection, or injury, can cause remodeling of vessels, manifesting as occlusions, kinks, tubulations, or other collateral formation of vessels. Pathway analysis identified gene sets of ECs (four sets), SMCs (2 sets), fibroblasts (1 set) and pericytes (1 set) which are the basic cell types composing vessel walls. Dysregulated response of vascular SMC can induce hypertension, and excessive proliferation of these cells contributes to CVD progression [85]. ECs dysfunction can lead to hyperpermeability, neurovascular decoupling and proinflammatory responses [7]. We identified a gene set for “human retinal fibroblasts” consistent with the fact that this cell type is the most common in connective tissue and involved in maintaining the extracellular matrix. Under stress, fibroblasts proliferate resulting in the accumulation of extracellular materials that ultimately limits elasticity [86]. In addition, we found enrichment in a gene set related to “mesangial cells”, which are kidney-specific pericyte cells. Retinal capillaries are composed of endothelial cells and pericytes. These contractile cells control blood flow in capillaries [87] and their function is inhibited under stress, such as in high glucose conditions typical in diabetes [88]. Therefore dysregulation of these gene sets has the potential to induce vessel remodeling under stress.

We identified genes directly involved in vessel remodeling. In particular, *FLT1* plays a role in the process of collateral vessel formation, which is a form of vascular remodeling in response to stress, such as hypoxia or hypertension [89]. *FLT1* is transcribed in several tissues including arteries and heart [71] and translated into VEGFR1. VEGFR1 is upregulated in response to micro-inflammation in the early stages of several vascular diseases [89]. In the retina, VEGFR1 is observed in ECs, SMCs, pericytes and RPE cells (which modulate fibroblast proliferation), and excess VEGFR1 contributes to vessel leakage and angiogenesis [89].

### Associations with diseases

We detected pleiotropic effects of tortuosity loci, which we showed to be independently associated with CAD, myocardial infarction, hypertension, diabetes, chronic lymphocytic leukemia, Alzheimer’s disease, myopia and glaucoma. We also found tortuosity genes to be involved in disease pathomechanisms. *ACTN4*, our top hit, was recently associated with vasorelaxation [90], a mechanism that can lead to hypertension when malfunctioning. The lead SNP in *ACTN4* tortuosity (rs1808382) is also independently associated with CAD [45]. *COL4A1* mutation has been reported as the cause of retinal arteriolar tortuosity [91] and cerebral small vessel disease [92] vessel leakage and hyperpermeability [93]. Fittingly, *COL4A2* also figured among our variants with pleiotropic effects on disease risk (see Table 4). Variants in the fetal genome near *FLT1* have been associated with preeclampsia [94], a condition of pregnant women presenting with hypertension and damage to the liver and kidneys, whose underlying mechanism involves abnormal formation of blood vessels in the placenta [95]. Retinal vessel modifications have been observed to precede clinical onset of preeclampsia and persist upto 12 months postpartum [96–98].

We elucidated causal links between tortuosity and disease risk factors by applying MR. Specifically, we established that elevated LDL exposure causally reduces arterial tortuosity. High LDL is known to cause the buildup of atherosclerotic plaque [99], which has been clinically linked to arterial tortuosity [100,101]. In fact, arteriosclerosis may make retinal arterial walls less flexible and thereby reduce their DF. We observed a *negative* causal effect of venous tortuosity on BMI, despite the known *positive* correlation between BMI and retinal tortuosity [102], suggesting that environmental factors may play a role in the relationship between BMI and vascular tortuosity.

### Limitations

This study was subject to the following limitations. First, we focused on the DF as a tortuosity measure, since the corresponding GWAS revealed many more significant loci, genes and pathways, as well as a higher heritability estimate in comparison to the alternative curvature-based tortuosity measures. These measures are more sensitive to local physiological vessel features, such as aneurysms or sharp bending (“kinks”), while DF only captures the total vessel elongation. Interestingly, the GWAS for these measures revealed several specific genes and pathways that were not significant in the DF analysis, which may be associated with pathologies manifesting as local disruptions in the microvascular network. Further work is needed to elucidate to what extent the stronger association signals for the DF are due to its robustness as a tortuosity measure or its quality to capture *total* vessel elongation as the most physiologically relevant trait. Second, due to the small size of our replication meta-cohort we only had enough power to replicate 4 of our 173 hits. Nevertheless, the effect sizes of SNPs and genes in the replication studies correlated strongly with those in the discovery cohort, providing independent evidence that they were not driven by any artifacts specific to the UK Biobank [52]. Moreover, our gene and pathway analyses indicate that these signals make biological sense. Finally, we did not adjust for spherical equivalent refractive error, which may have confounded our measurements to some degree. Investigating the impact of this and other corrections on the heritability estimates could shed more light on their usefulness.

### Conclusion

This study exploits advanced automated image processing and deep learning to characterize different *vessel type specific* retinal tortuosity measures from >100k fundus images to conduct a high-powered GWAS on this trait. The resulting significant association signals for 175 genetic loci allowed us to estimate the heritability of tortuosity, identify genes, annotated gene-sets and tissues relevant for this trait, and reveal pleiotropic links with and causal effects to or from disease-related traits. This range of analyses was largely possible due to the large data set made available by the UK Biobank, in terms of both genetic analysis and the high-dimensional data available from fundus images. Combining these data provided novel insights into the genetic architecture of retinal tortuosity, the associated morphological features, the potential disease processes, and the identification of candidate targets for ocular and systemic treatments.

## Data Availability

The data are made available by the UK Biobank Consortium.

## Nonstandard Abbreviations and Acronyms

BMI: body mass index
BRB: blood-retina barrier
CAD: coronary artery disease
CVD: cardiovascular diseases
DBP: diastolic blood pressure
DF: distance factor
DVT: deep vein thrombosis
EC: endothelial cells
GO: gene ontology
GWAS: genome-wide association study
LD: linkage disequilibrium
LDL: low-density lipoprotein
SBP: systolic blood pressure
SMC: smooth muscle cell
SNP: single nucleotide polymorphism
VEGF: vascular endothelial growth factor

## ACKNOWLEDGEMENTS

This work was conducted using the UK Biobank (application ID 43805) and SKIPOGH. Thanks to Micha Hersch for inspiring this project, to the UK Biobank team for their support and responsiveness, and to all UK Biobank participants for sharing their personal data. We also thank aSciStance Ltd for their help in revising the manuscript.

## AUTHOR CONTRIBUTIONS

MT and SB designed the study. MT and MJB performed QC on the raw images. MT and SOV extracted tortuosity measurements from the image data (UK Biobank, *OphalmosLaus* and SKIPOGH). MJB performed classification of arteries and veins. MT carried out the median DF tortuosity GWAS, with the guidance of SB, NM and EP. SOV carried out the tortuosity GWASs based on alternative measures with input from MT. MJB and SOV carried out gene and pathway scoring with the guidance of DK. ALB performed LD Score Regression analysis. SOV evaluated the correlation between different tortuosity measures and their impact on genetic associations. MT and TC performed the replication analysis in SKIPOGH with input from MB. HA, LK, RS and CB provided ophthalmological expertise and manually annotated the raw image data. MT, CB and SB lead the writing of the manuscript with contributions from all other authors.

## SUPPLEMENTAL

Supplemental Methods (Supplemental Text 1–5)

Supplemental Results (Supplemental Text 6–13)

Supplemental Datasets 1–3, 4A–C, 5, 6A–C, 7A–C

## SUPPLEMENTAL MATERIAL

## SUPPLEMENTAL DATASET

[Supplemental Dataset 1 - 175 lead SNPs](https://docs.google.com/spreadsheets/d/1mhy8mRZs_bUdFyL331dPPU_N0-mBPAQ9LsHMCzPFGfE/edit?usp=sharing)

[Supplemental Dataset 2 - overlap with GWAS Catalog (SNPs in LD)](https://docs.google.com/spreadsheets/d/1wYTp7pweURdA5-MINn5LWXQ7oT_4Z1e8E56kAgDkfbc/edit?usp=sharing)

[Supplemental Dataset 3 - overlap with GWAS Catalog (SNPs with matching rsIDs)](https://docs.google.com/spreadsheets/d/1dTh9yhHiRfQqBzPkUoHazvo4vSvBEJX9gL_bFGzfoR0/edit?usp=sharing)

[Supplemental Dataset 4A - significant SNPs (combined)](https://docs.google.com/spreadsheets/d/1PUSbmK_snLIdW4wtx7YqEUhrCPlWRH6fLfsUE4T-3dg/edit?usp=sharing)

[Supplemental Dataset 4B - significant SNPs (artery)](https://docs.google.com/spreadsheets/d/1ei2pogiEswGmz36-dXvMa61Kd4vFvVDVkCcIuqG43po/edit?usp=sharing)

[Supplemental Dataset 4C - significant SNPs (vein)](https://docs.google.com/spreadsheets/d/1Q0tyyNL7lOfLIqX4nVHbIJdJpGFrKCV3M8MakI2zaa8/edit?usp=sharing)

[Supplemental Dataset 5 - replication of lead SNPs](https://docs.google.com/spreadsheets/d/1M5u83p-7ljei_TLbHrgnRU1ays-xYv4QExD-8GOKro8/edit?usp=sharing)

[Supplemental Dataset 6A - gene scores (combined)](https://docs.google.com/spreadsheets/d/19_1skGdPyYilA_bOGNaWrdezrCRauxTGGuFQyWoABKc/edit?usp=sharing)

[Supplemental Dataset 6B - gene scores (artery)](https://docs.google.com/spreadsheets/d/1koU44Ca6jRZPADqk8cWGkgl0NngQGG5MzmzQBQX673o/edit?usp=sharing)

[Supplemental Dataset 6C - gene scores (vein)](https://docs.google.com/spreadsheets/d/1T4jqBUnd5dxqsyrVC3jzFhWlRVPqTP9jGJtqLAIPETc/edit?usp=sharing)

[Supplemental Dataset 7A - gene set enrichment (combined)](https://docs.google.com/spreadsheets/d/14iuAwCYAY49CwaqZqigXPBiP2iBgoZnYO7zbyMktnpw/edit?usp=sharing)

[Supplemental Dataset 7B - gene set enrichment (artery)](https://docs.google.com/spreadsheets/d/1vLyStn6tPZ4Prbg93IQMJDM3vl-qnGE29a_teODlegU/edit?usp=sharing)

[Supplemental Dataset 7C - gene set enrichment (vein)](https://docs.google.com/spreadsheets/d/1MakdTwy7JKr3L3E_WyDCePeZj4fJbRzRTt9Ib7NjrtQ/edit?usp=sharing)

## SUPPLEMENTAL METHODS

### Text 1: Quality Control in tortuosity measurements

The quality control (QC) procedure is performed during data extraction. The objective is to remove (i) lower quality images and (ii) images containing artifacts. QC relies on thresholding being applied to two distributions: 1) Distribution of the total number of equally spaced diameters that were fitted to the vessels of each eye. Images between with values 11 000 and 20 000 passed QC. 2) Distribution of the number of vessels contained in each eye. Images with values between 100 and 250 passed QC. By visual inspection, we fine tuned these thresholds to discriminate (i) low-quality images that were too dark, too light, out of focus (lower-bound thresholds), or (ii) images that contained spurious vessels, i.e., artifacts of the picture that were being erroneously segmented as vessels (higher-bound thresholds). Images that pass QC (roughly two out of three) are further processed to extract several tortuosity measures.

### Text 2: DF and other tortuosity measures

#### Definition of DF

We consider a vessel as a curve in a two dimensional space on the interval [t_0_, t_1_]. For a given curve there are different tortuosity measures: The simplest one, the “Distance Factor” (DF), is calculated as the total arc length over total chord length. This measure computes the tortuosity of the segment by examining how long the curve is relative to its chord length.

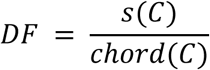

Let *s*(*C*) be the arc length of the curve C, and *chord*(*C*) its chord length:

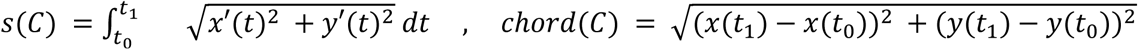

where the prime denotes derivation with respect to *t*.

#### Definitions of six, alternative, curvature-based measures

A first curvature-based tortuosity measure is obtained as the integral over the absolute value of curvature along the entire curvature:

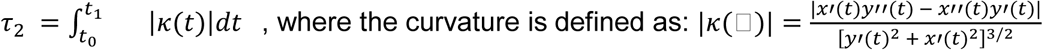

This is equivalent to the inverse curvature radius *R(t)*, i.e. the radius of a circle that is tangent to the curve at t: |*κ*| = *1/R*.

A second tortuosity measure is the integral over the curvature squared along the entire curve:

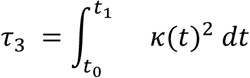

Finally, four tortuosity measures arise from normalizing and □_3_ by division through either the curve or the arc length:

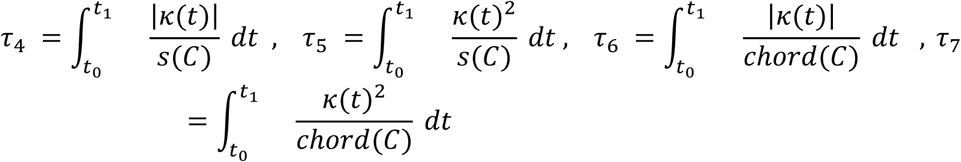

We note that, by definition, □_2_ and □_3_ depend on the length of the curve (with non-zero curvature). In the case of vessels with constant curvature radius: □_2_= *s(C)/R* and □_*3*_*= s(C)/R*^*2*^. Our analysis shows that □_2_ differs from other measures the most, followed by □_3_, while □_4-7_ are indeed quite similar to each other (see Supplemental Figure 1)

#### PCA of DF and curvature-based measurements

**Supplemental Figure 1.**
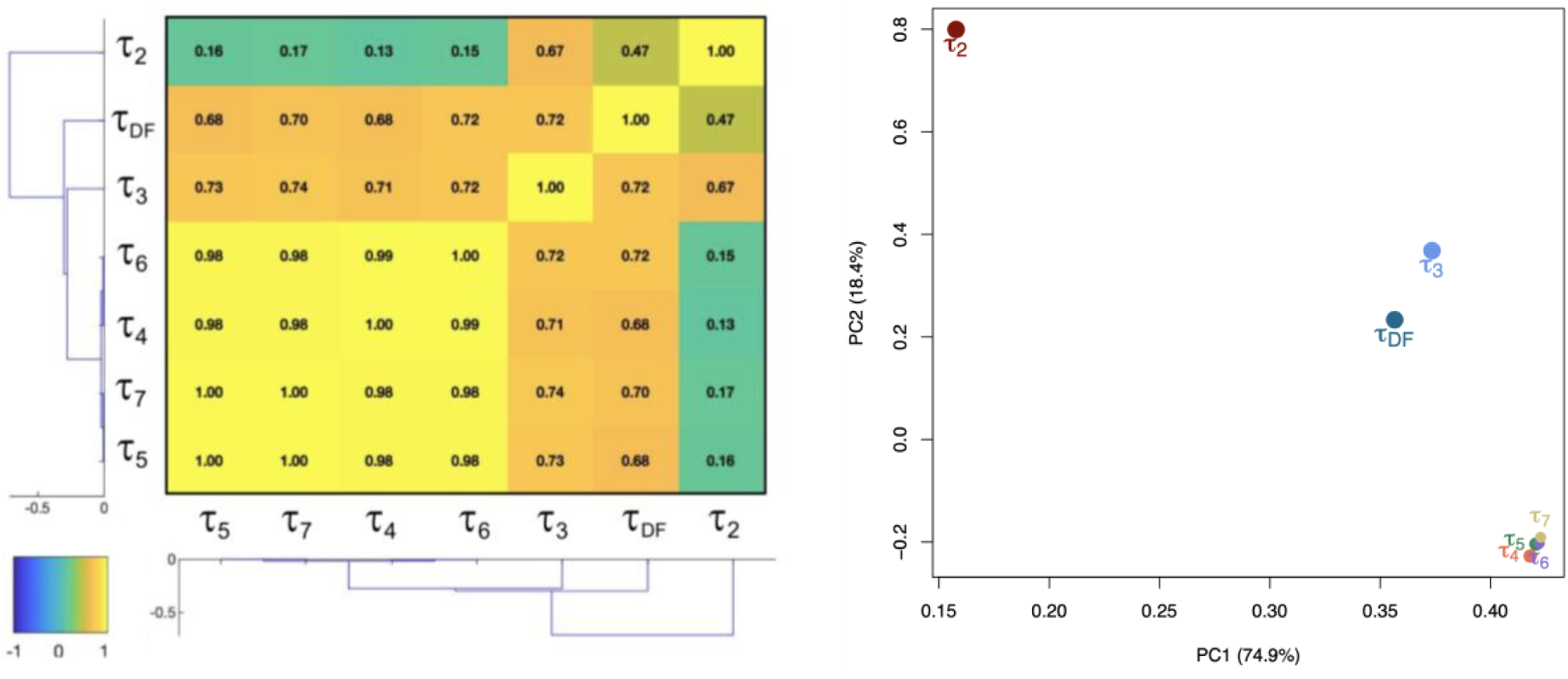
Dimensionality reduction on tortuosity measurements across 62 751 individuals. Left: Hierarchical clustering of the pairwise correlations-matrix. Correlations are measured using the Pearson correlation coefficient. Right: We performed dimensionality reduction using a Principal Component Analysis (PCA) approach and plotted PC2 against PC1.

#### Heritability of DF and curvature-based measurements

**Supplemental Table 1.**
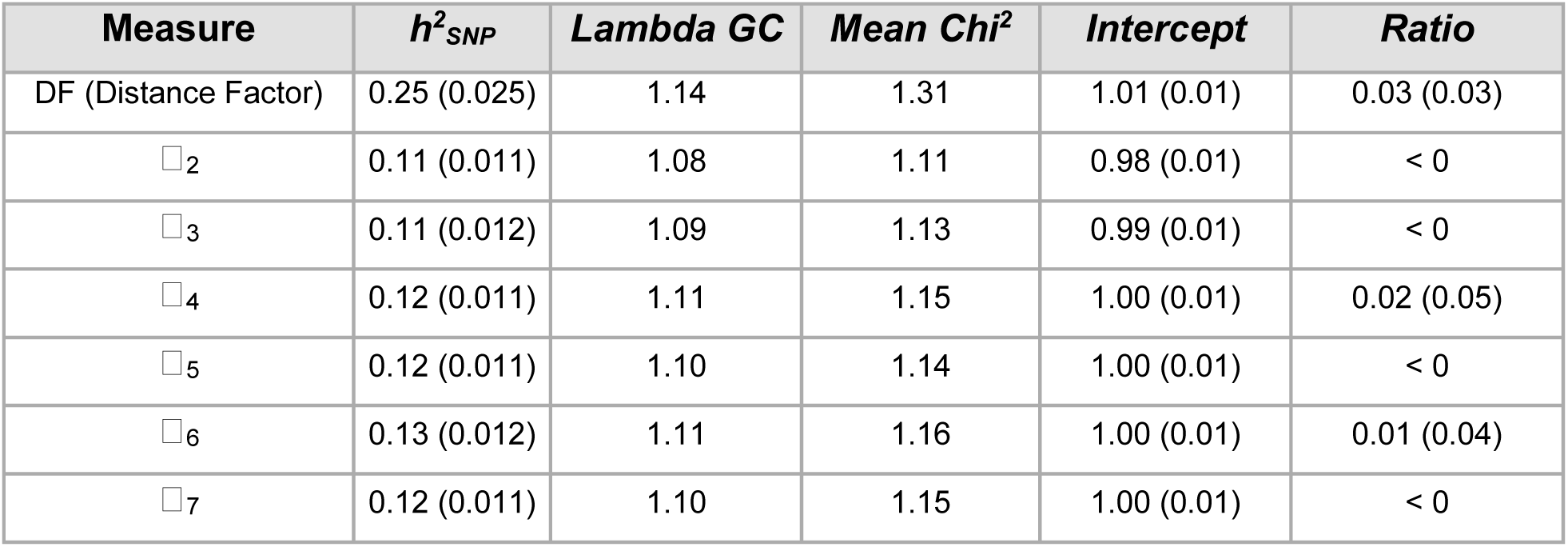
SNP-based heritability of alternative tortuosity measures. h^2^_SNP_: is the portion of phenotypic variance cumulatively explained by the SNPs. Traits defined by the six alternative tortuosity measures were less heritable than the one defined by the Distance Factor. *Lambda GC* is the measure of inflation (it measures the effect of confounding and polygenicity acting on the trait). *Intercept* is the LD Score regression intercept (values close to 1 indicates little influence of confounders, mostly of population stratification). *Ratio* is the ratio of the proportion of the inflation in the *Mean Chi*^*2*^ that is not due to polygenicity (a ratio close to, or smaller than, 0 is desirable as it indicates low inflation from population stratification). SE are given in parentheses.

#### Enrichment analysis of curvature-based measurement associations

The alternative tortuosity measures have less significant SNPs, genes and pathways overall compared to the distance factor, but harbor few unique pathways, among which is a pathway, in □_6,_ called “abnormal cardiac ventricle morphology” (-log_10_ p=5.8) from the human phenotype oncology (HP) group. Being the gene scores of them highly significant in our analysis.

On the phenotypic level, we found that tau 2 (total curvature) is least similar to all the others, that tau 3 (total squared curvature) is comparably similar to the DF, and that tau 4-7 (average curvature) form a distinct cluster of similar measurements.

Here, despite all alternative measures having significantly lower SNP-wise heritability than the DF, we found eighteen genes (Figure 2a) and four pathways (Figure 2b) specific to them, i.e. not present in the DF. Among them are genes with potentially relevant annotations, as described by *GeneCards* [24]: OCA2 (Oculocutaneous Albinism 2), the overall top hit, is a determinant of eye color, and associated with albinism. TRIOBP, the most significant specific hit in tau 2, is a structural protein binding to F-actin, and acts as a stabilizer of the cytoskeleton. LGALS1 is implicated in modulation of cell-cell and cell-matrix interactions, and thus a structural protein that might affect, among other things, vessel bendiness. GLIS3 is a zinc-finger protein that plays a role in eye development.

We also identified a cluster of four genes (LRIT1, LRIT2, CDHR1, RGR) on locus 10q23 (Supplemental Figure 2c) through STRING [25], all of which have been associated with eye disorders. RGR has been associated with Retinitis pigmentosa. LRIT1, LRIT2 and CDHR1 have been associated with nanophthalmos, a developmental eye disorder characterized by small eyes. Molecularly, CDHR1 is a calcium-dependent cell adhesion molecule expressed in blood vessels, and thus likely to affect their morphology.

Two out of the four pathways both specific to the average curvature measures, are plausibly related to vascular changes. The first, GO_CELL_JUNCTION_ORGANISATION, is a GO set of genes influencing the tightness of connection between neighboring cells. The second, HP_ABNORMAL_CARDIAC_VENTRICLE_MORPHOLOGY, Human Phenotype Ontology set of genes associated with disease-related abnormalities in cardiovascular tissue. This pathway has three genes strongly driving the signal: 1) RYR1, a sarcoplasmic reticulum calcium release channel, 2) MYOZ2, a sarcomeric protein involved in calcium-dependent signal transduction, and 3) FADD, an apoptotic adapter molecule. Interestingly, no pathways were specific to the most dissimilar measure, tau 2.

In summary, first, this confirms two observations we made on the phenotype level: 1) tau 2 with twelve unique gene hits is the most dissimilar from the others, and 2) tau 4-7 are almost identical and can probably be treated as one measure. Second, we identified relevant genes and pathways specific to these measures, indicating that they may capture disease-relevant vascular changes the DF is not sensitive to.

**Supplemental Figure 2.**
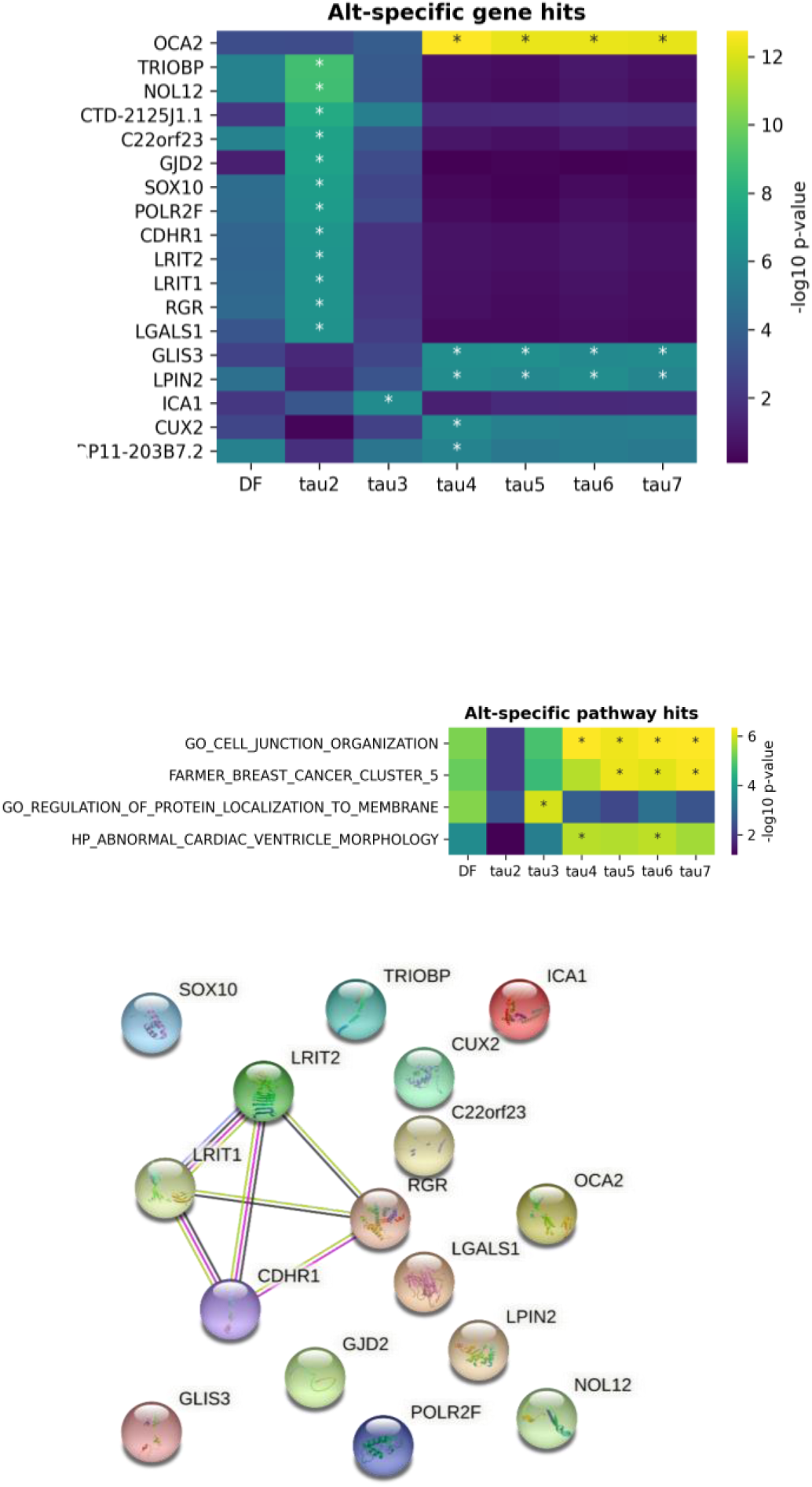
Genes and pathways specific to alternative tortuosity measures. We use a Bonferroni-corrected -log10 p-value significance threshold of 5.71 for genes, and 5.79 for pathways. Significant hits are marked with an asterisk (*). The tables are ordered by maximal significance. a. All the genes and pathways that are significant in one of the alternative tortuosity measures, but not in the distance factor (DF), are displayed. There are twelve genes specific to tau 2, one to tau 3, and five to the very similar measures tau 4-7. b. Four pathways are specific to the alternative tortuosity measurements: one in tau 3, three in tau 4-7, but none were found in tau 2. They contain 577, 10, 170, and 394 scored genes respectively. c. STRING [25] cluster of four genes (LRIT1, LRIT2, CDHR1, RGR), all of which reside on locus 10q23, and have been associated with eye pathologies.

### Text 3: Distribution of DF

#### DF tortuosity across cohorts

**Supplemental Figure 3.**
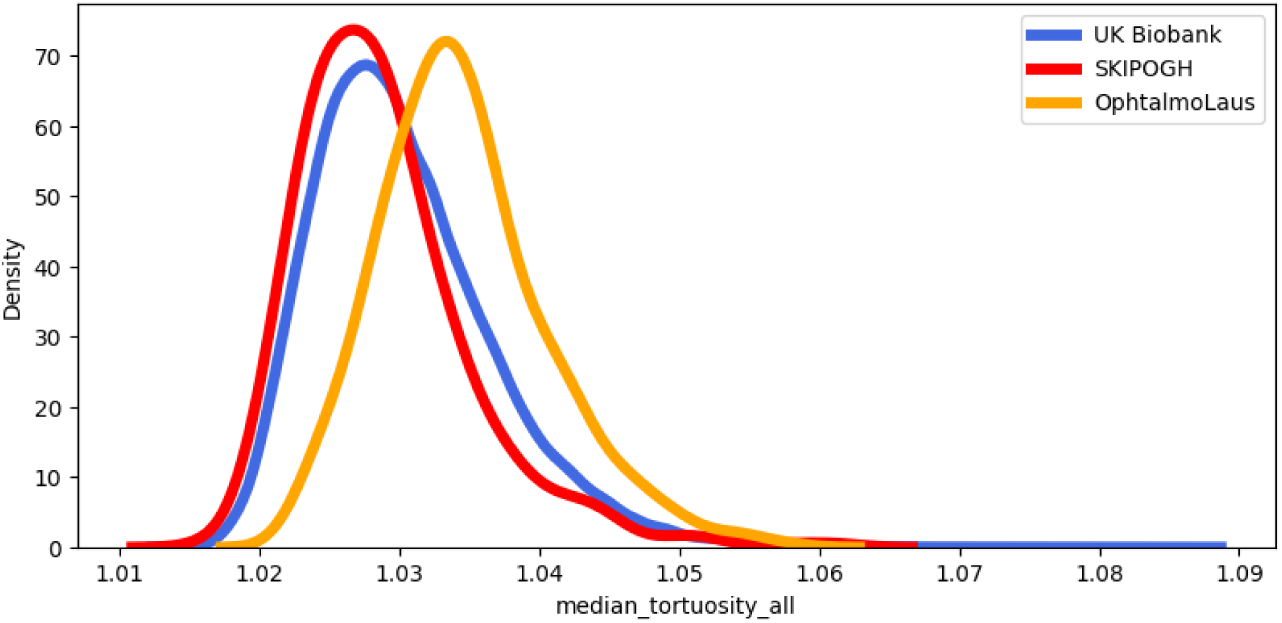
Distribution of DF tortuosity across cohorts. UK Biobank mean±SD = 1.030±6.5×10^−3^; SKIPOGH mean±SD = 1.029±6.2×10^−3^; *OphtalmoLaus* mean±SD=1.034±6.0×10^−3^.

#### Stratified DF analysis: sex, age and vessel type

**Supplemental Figure 4.**
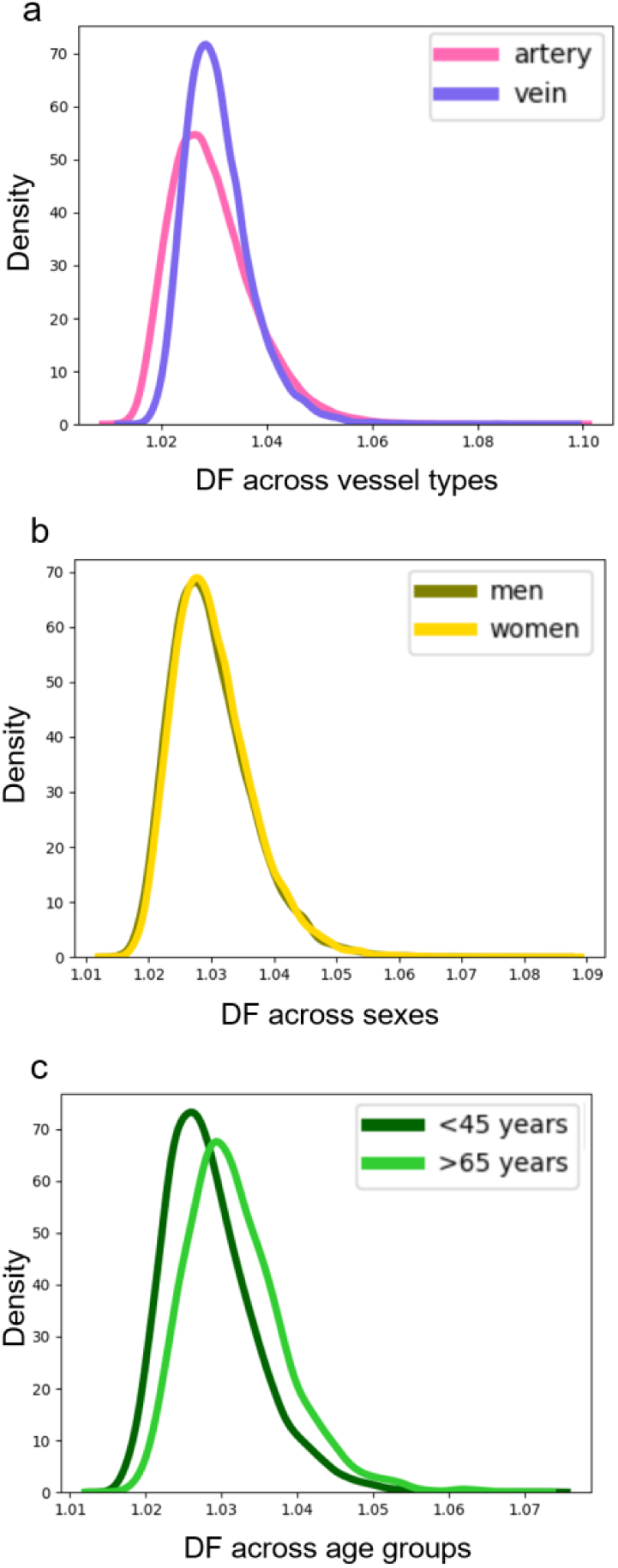
Median DF between age groups, sexes and vessel types in the Uk Biobank. **a**. distribution in arteries (DF = 1.029) and veins (DF = 1.030); Cohen’s d = 0.13, *p =* 9×10^−142^. **b**, distribution in men (DF = 1.0300) and women (DF = 1.0304); Cohen’s d = 0.049, *p =* 9×10^−10^. **c**, median DF increases as a function of age. In the youngest decile (<45 years) DF = 1.028, while in the oldest decile (>65 years) DF = 1.031. Cohen’s d = 0.49, *p =* 1×10^−195^.

### Text 4: Deep Learning classification of arteries and veins

#### Accuracy of vessel type classification

LWNET [2] converts raw RGB fundus images into a categorical image of three categories, which are defined by the following RGB values: 1) artery: red (255,0,0), 2) vein: blue (0,0,255) and 3) background: black (0,0,0) (Supplemental Figure 5a).

We used LWNET to perform automatic artery-vein classification on our 44 ground truth images (fundus images for which we have manual annotation by ophthalmologist HA), and subsequently extracted the resulting categories for vessel segment centerlines, which we previously computed using ARIA software. For all the centerlines of the 44 images we computed a segment score based on individual centerline pixel classifications as follows:

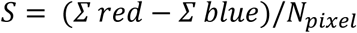

A ROC curve and its derived AUC measure was computed on the resulting vector of segment scores using logistic regression. This resulted in AUC=0.93. Accuracy was computed simply as:

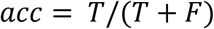

where *T* is the number of correctly categorized segments, and *F* the number of wrongly categorized segments. We chose not to censor any segments, and called classification based on the simple rule that vessels with S>0 were called arteries and vessels with S<0 were called veins. This resulted in *acc* = 0.88.

**Supplemental Figure 5.**
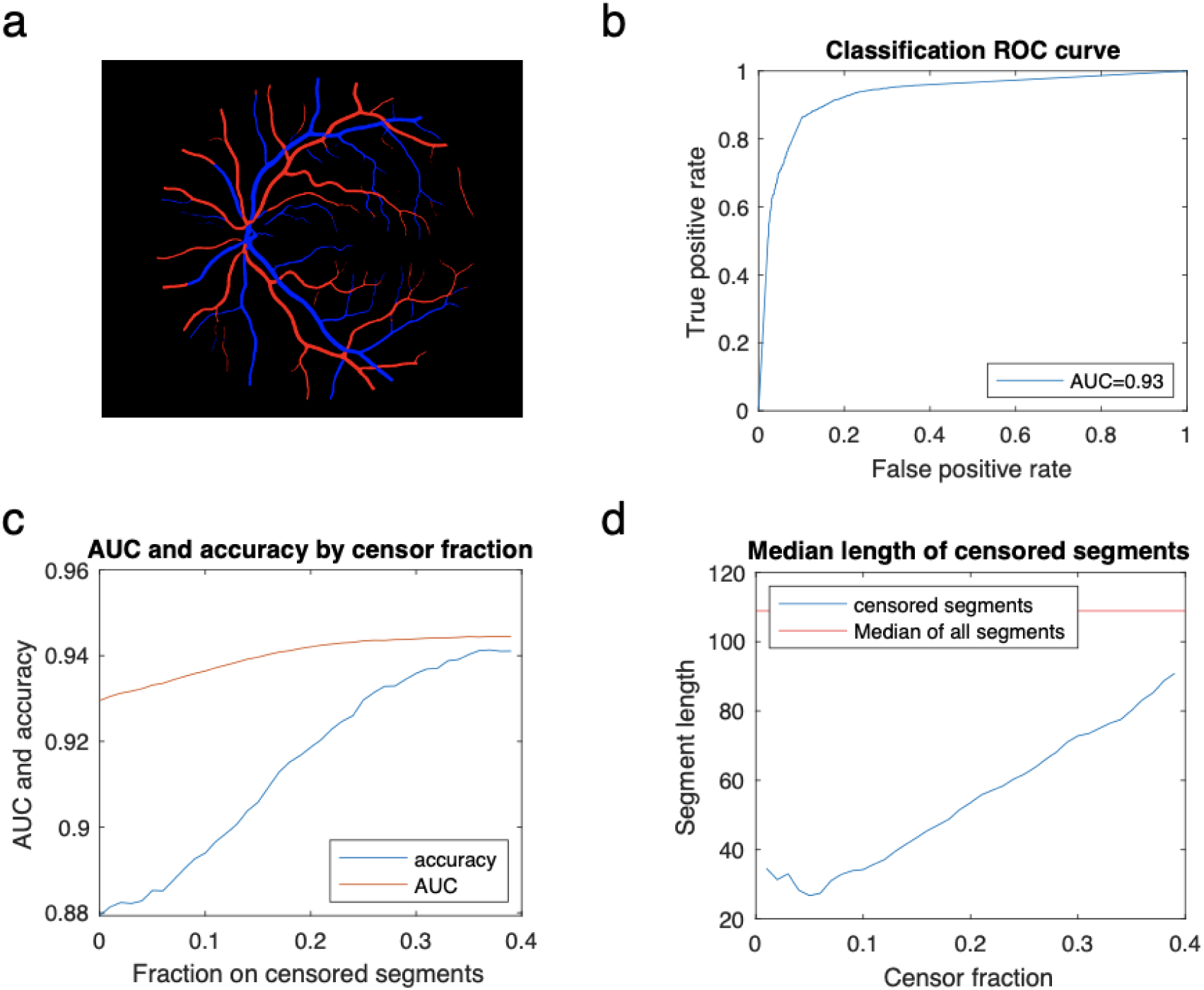
AUC and accuracy of artery-vein classification. a) LWNET output image consists of 3 categories: artery, vein and background. b) ROC curve when not censoring any segments, resulting in AUC=0.93 and accuracy=0.88. c) AUC and accuracy could be further improved by censoring the most unclearly classified vessel segments. d) Median length of censored segments is significantly shorter than the overall median length, which could be the reason why we found decreased heritability in our traits in the censoring approach.

#### Censoring unclearly classified vessels

Performance could be further increased by removing segments with more uncertain scores (i.e. close to zero). We increasingly removed the vessel segments with the lowest absolute score, and measured how this influenced AUC and accuracy based on the remaining segments. We found that, while AUC shows only moderate increase, accuracy increased from 0.88, without censoring, to almost 0.94 when removing a third of the segments (Supplemental Figure 5c). However, we also found that by doing this we predominantly removed shorter segments (see Supplemental Figure 5d).

We then ran a GWAS based on only the approximately ⅔ most confidently scored segments in the UK Biobank, and found a doubling of the number of Genome-Wide-significant hits for arteries (+117%) (the amount of signal for veins was, on the contrary, not affected). Further analysis, though, showed an inflation in the Q-Q plot of the artery-specific GWAS based on high-confidence vessels only. This was confirmed by analysis of the parameters of the LD Score Regression, which indicated a loss in the ability of the results to explain (SNP)-based heritability of the trait (h_SNP_^2^ dropped from 0.25 to 0.11), coupled with genomic inflation (intercept had increased from 1.01 to 1.93). For these reasons, all vessels identified as arteries or veins were used in the respective vessel-type-specific analysis, as selecting vessels with the highest identification score had brought marginal improvement to the already high AUC at the price of introducing a bias.

#### GWAS with random vessel type calling

We estimated the independence of the signals arising from the above-described classification of arteries and veins as follows: we modified the pipeline to perform random calling of arteries and veins (by shuffling the vector of artery and veins scores computed for each eye). We then compared the similarity in the signal between two random vessel-type GWAS: we clearly show that the effect sizes (which are significant in at least one of the two GWAS) are nearly identical (*r*=0.99, *p*=4·10^−82^) when the vessel type calling is random. By comparison, the effect sizes are much less coupled (*r*=0.76, *p*=1·10^−20^) in the artery- and vein-specific GWAS based on the vessel type calling procedure that has been described

**Supplemental Figure 6.**
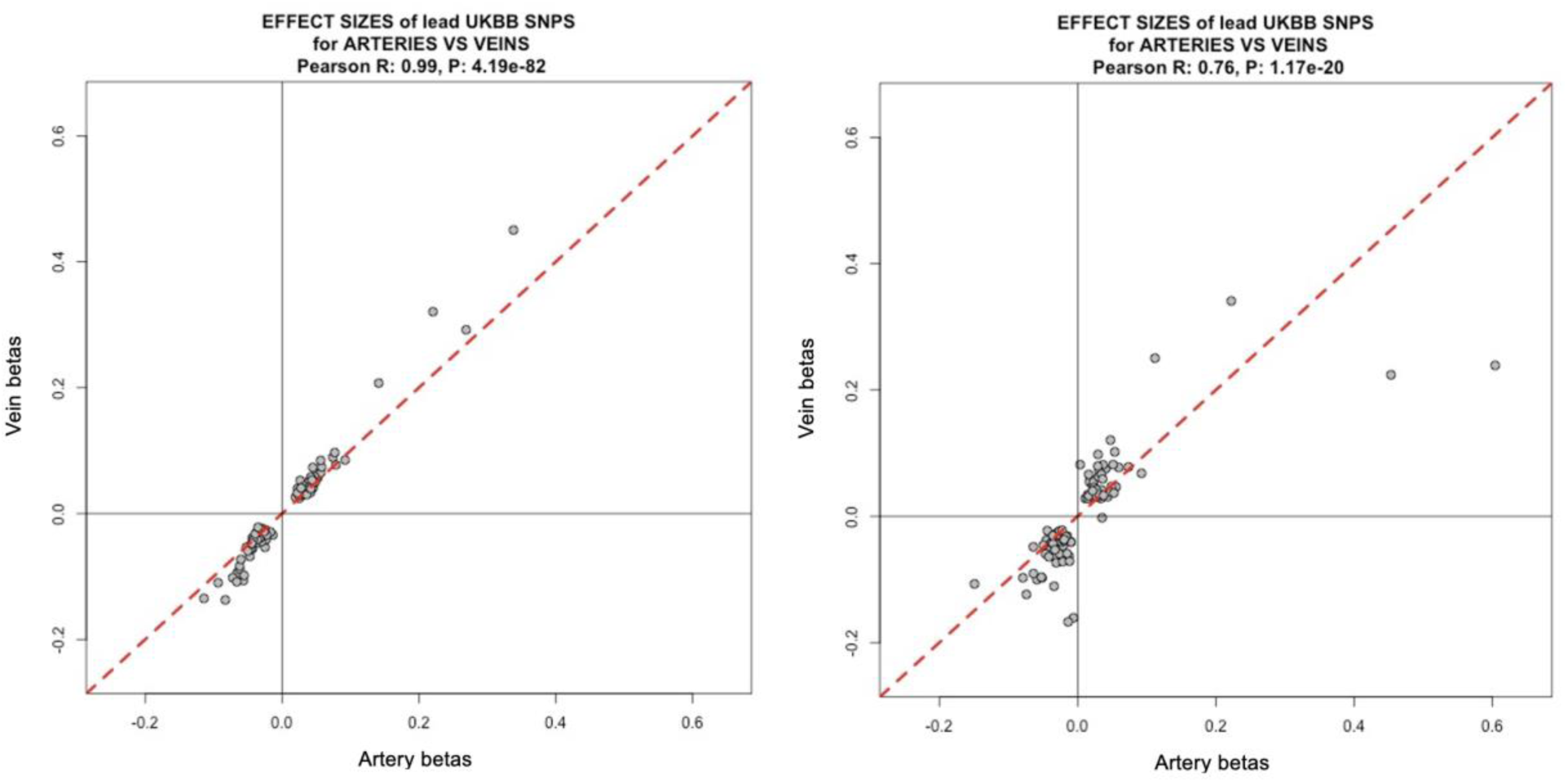
Correlation in the effect sizes from arteries and veins. To support the effectiveness of our vessel type calling procedure, we performed random calling of arteries and veins (by shuffling the vector of artery and veins scores computed for each eye). We then performed two random vessel-type GWAS (left) and compared the results with the artery and vein-specific GWAS (right): we clearly show that the effect sizes (which are significant in at least one of the two GWAS) are less coupled when the vessel type is not random.

### Text 5: Replication Analysis

#### Correlation of effect sizes in the meta-cohort

Of the 136 SNP shared between the discovery and replication studies, the sign of the effect sizes was concordant in 90 (binomial test p = 5.0×10-5). We observed a Pearson correlation of *r* = 0.53 (*p* = 1.2×10^−11^) between the effect size estimates in the two studies. When some outliers are removed, the correlation drops from *r* = 0.53 to *r* = 0.36, remaining highly significant.

**Supplemental Figure 7.**
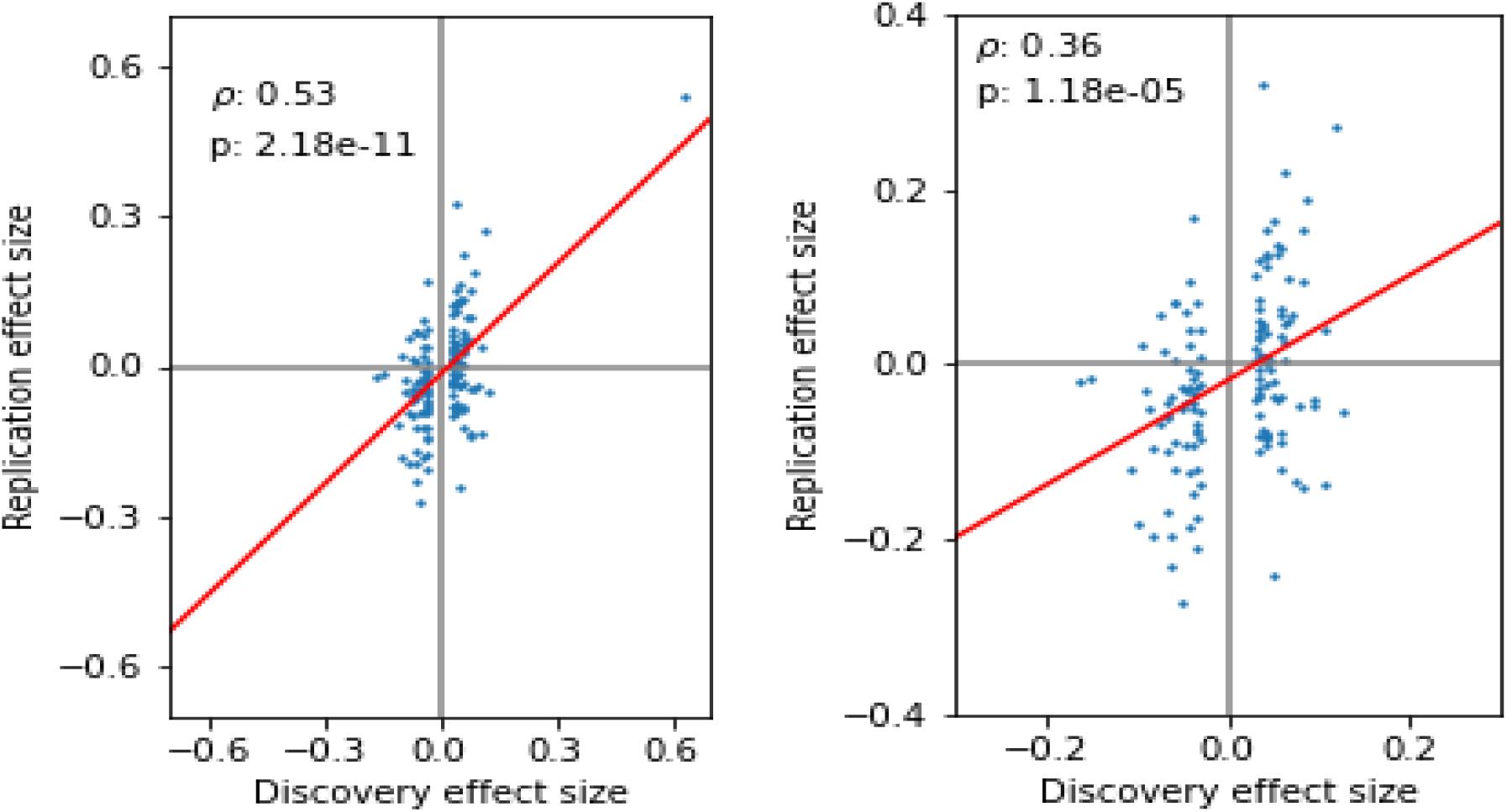
Extended plot for correlation of effect sizes between discovery and replication cohort. Left: correlation of effect sizes in the discovery and replication cohort as shown in the main text of the manuscript. Right: same plot without considering the outlier. We removed one outlier in the top-right quadrant, corresponding to rs187691758. This shows the robustness of the result: even though the Pearson correlation dropped from *r*=0.53 to *r*=0.36, it remained highly significant (p=1.18·10^−-5^). Out of the 135 remaining SNPs, 89 shared the sign in effect size (binomial test p=2.94·10^−6^).

#### Replication of hits in the meta-cohort

We performed a meta-analysis of the two cohorts OphtalmoLaus (N=514) and SKIPOGH (N=397), using both a fixed-effects model (see below) and a random-effects model [3]. For each SNP, the fixed-effects model computes meta-values of the standard error (SE) and effect size summary statistics, with the meta effect size being a weighted average of individual effect sizes, weighted by their corresponding inverse SE:

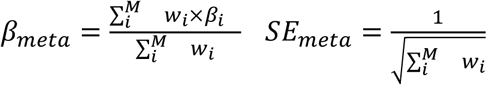 where 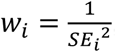, and M the number of cohorts

The resulting SNP P-value is then given by a two-tailed t-statistic

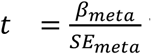 with 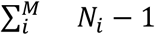 1 degrees of freedom (911 in our case)

The following two figures present the results from both meta-analyses, and the subsequent gene and pathway scoring based on their results. All replication is performed on the combined-vessel median distance factor phenotype.

**Supplemental Figure 8.**
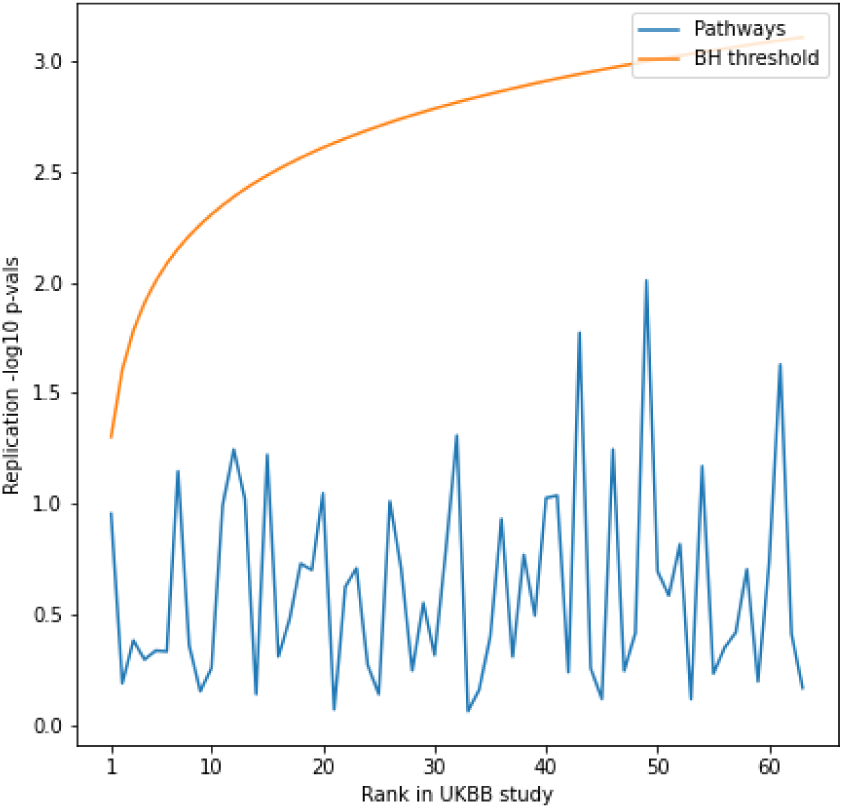
Replication meta-analysis with fixed-effects. Meta-GWAS summary statistics were obtained using the **fixed-effects** model described above. SNP-level and gene-level summary statistics were reported in the main text. SNP-level summary statistics were aggregated into pathway-level p-values using *PascalX*. Benjamini-Hochberg (BH) procedure on significant pathways from the discovery cohort doesn’t replicate any hits in the replication meta-cohort (SKIPOGH + OphtalmoLaus)

**Supplemental Figure 9.**
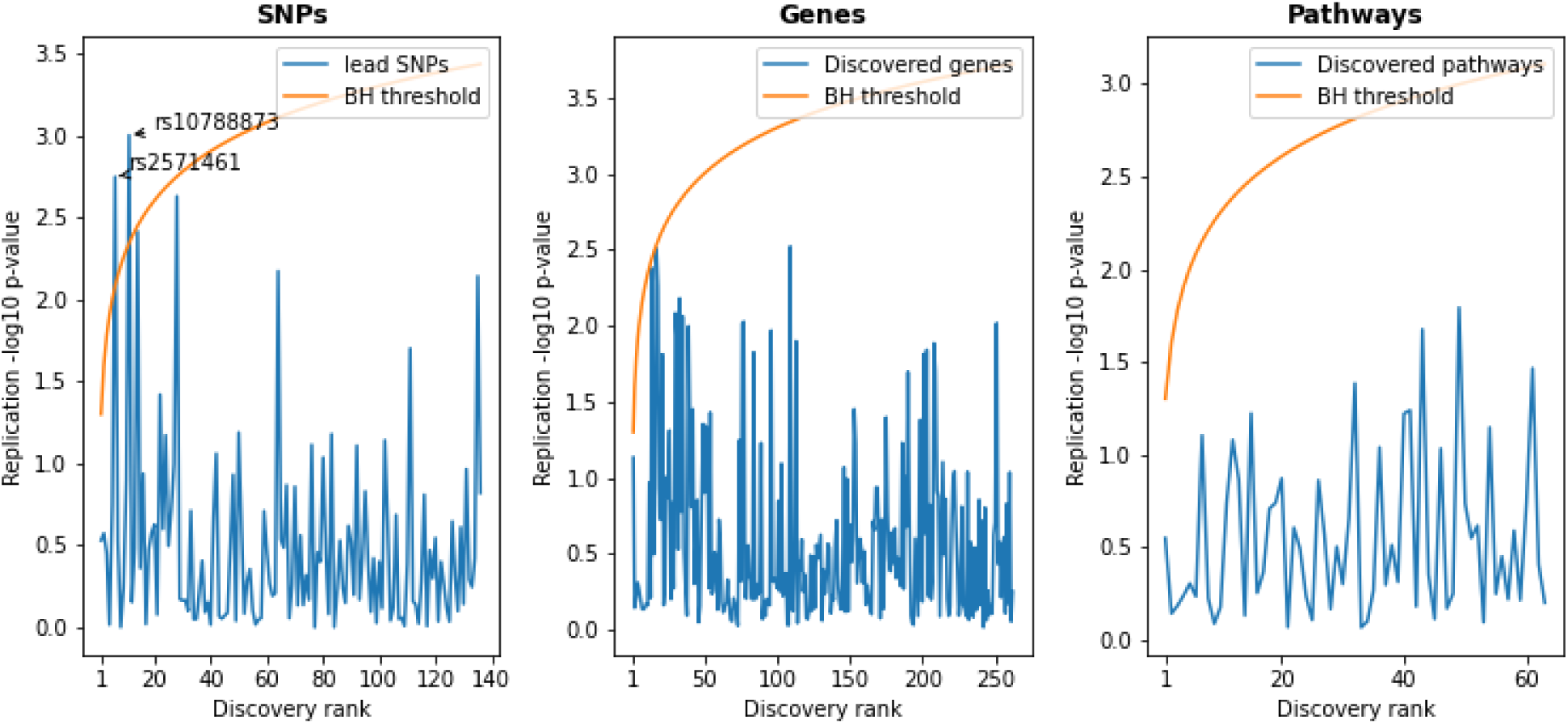
Replication meta-analysis with random-effects. Meta-GWAS summary statistics were obtained using the **random-effects** meta-analysis tool RE2C. Left: Benjamini-Hochberg (BH) procedure on lead SNPs discovered in the UK Biobank yields 4 hits in the replication cohort. Center and Right: No discovered genes and pathways replicate in the meta-cohort. SNP-wise p-values were aggregated into gene and pathway scores using PascalX.

## SUPPLEMENTAL RESULTS

### Text 6: Baseline Characteristics

Across the analyzed individuals, mean±SD age = 56±8 years; 35 098 females at birth (54%); 4 618 smokers (7%), mean±SD BMI = 27±5 kg/m2, mean±SD SBP = 140±20 mmHg, mean±SD DBP = 82±11 mmHg. 54 343 (94%) self-reported ethnicity as White, 1 243 (2.1%) as Asian, 962 (1.7%) as Back, 373 (0.6%) as Mixed, 175 (0.3%) as Chinese, and 521 (0.9%) as Other.

Among the participants for which at least one retinal fundus image was available, 2 644 had been diagnosed with type 2 diabetes, 1 448 with angina, 1 077 with myocardial infarction, 1 072 with deep-vein thrombosis (i.e., blood clot in the leg), 750 with stroke, 8 797 lived with stage 2 hypertension (i.e., automated reading of blood pressure >90 mmHg diastolic or >140 mmHg systolic); among the participants for which at least one fundus image was available, 2 644 had been diagnosed with type 2 diabetes, 1 448 with angina, 1 077 with myocardial infarction, 1 072 with deep-vein thrombosis (i.e., blood clot in the leg), 750 with stroke, 8 797 lived with stage 2 hypertension (i.e., automated reading of blood pressure >90 mmHg diastolic or >140 mmHg systolic).

### Text 7: Correlation with disease status

We built a logistic regression classifier and found retinal vein tortuosity to have predictive power over disease outcome: angina AUC 55.2%, heart attack AUC 53.4%, stroke AUC 54.6%, Deep Vein Thrombosis (DVT) AUC 53.3% and hypertension AUC 56.6%. To determine whether retinal tortuosity might be used as an independent biomarker for CVD, we trained logistic regression models with known risk factors: age, sex, SBP and smoking (pack years): angina AUC 76.2%, heart attack AUC 80.6%, stroke AUC 69.3%, DVT 61.8%, The same procedure was applied to the prediction of hypertension, but without using SBP as a risk factor, resulting in AUC 75.4%. These risk factors models did not significantly increase in performance by adding any of the median vessel tortuosity measures: we conclude that, despite associations with CVD outcome, retinal tortuosity does not represent an increased health risk after correcting for known risk factors. The analysis was performed on all vessels, then repeated only on veins and only on arteries. Results varied slightly. To illustrate this, we show the distributions of median vessel tortuosity in hypertensive patients vs. controls.

**Supplemental Figure 10.**
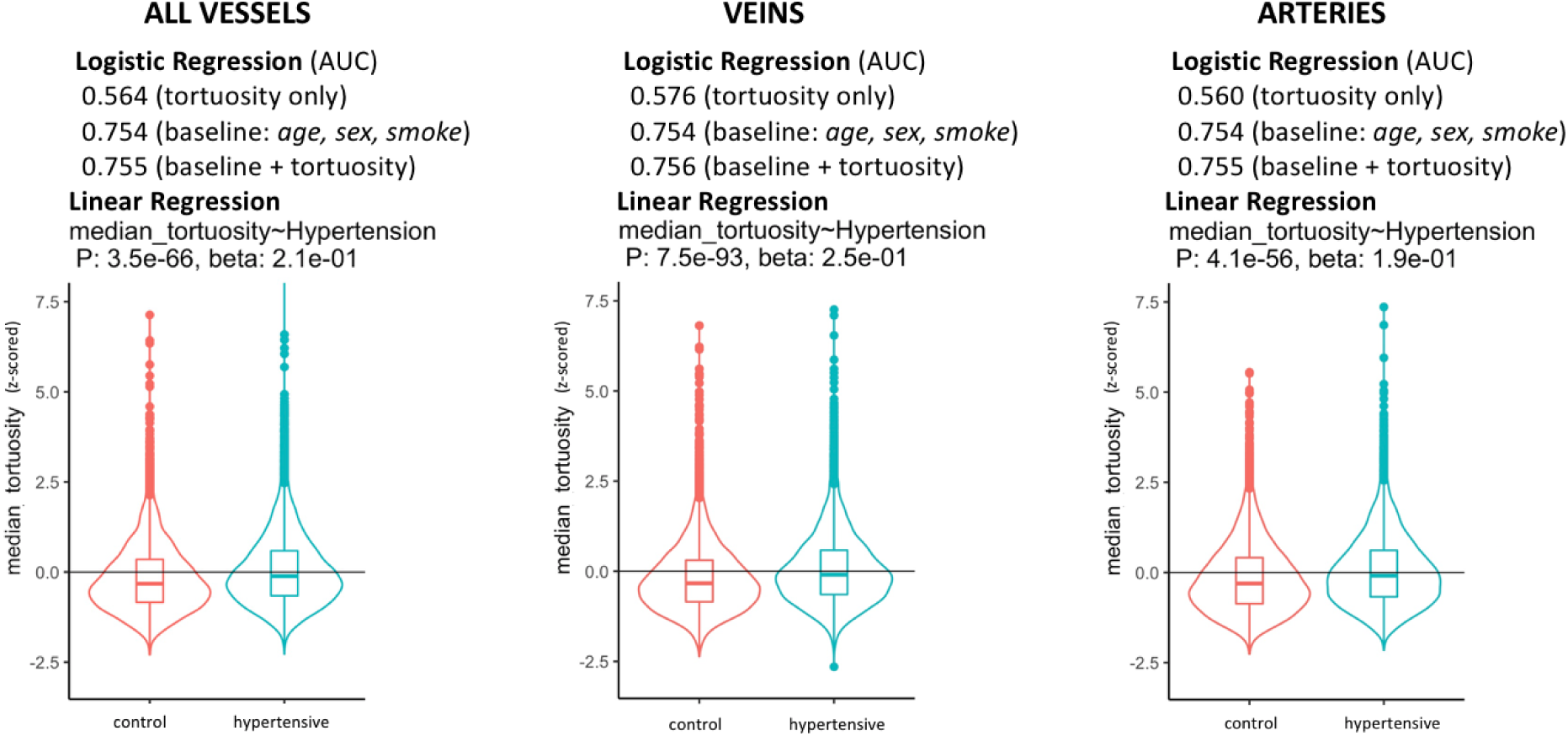
Predictive power of median tortuosity over hypertension. Effects and p-values are calculated using linear regression. AUC refers to logistic regression.

### Text 8: Replication of known hits

We replicated two known associations. We failed to replicate a third, for which association was controversial (it was only marginally significant in its discovery cohort and had failed replication in the independent cohort of the study that originally proposed it).

**Supplemental Table 2.** Known associations with retinal vessel tortuosity. Three SNPs known to associate with a phenotype related to retinal vessel tortuosity in the literature[4]. Details can be found in the list of all statistically significant SNPs (see Supplemental Dataset 1A/1B/1C).

| RSID | REPLICATED<br>in our GWAS | REPLICATED<br>in its own study | POSITION | GENE(S) | trait |
| --- | --- | --- | --- | --- | --- |
| rs1808382 | yes | yes | 19: 38.7 Kb | ACTN4 / CAPN12 | retinal venular tortuosity |
| rs7991229 | yes | yes | 13: 111.1 Kb | COL4A2 | retinal arteriolar tortuosity |
| rs73157566 | no | no | 12: 129.5 Kb | NLRP9P1 | retinal venular tortuosity |

**Supplemental Figure 11.**
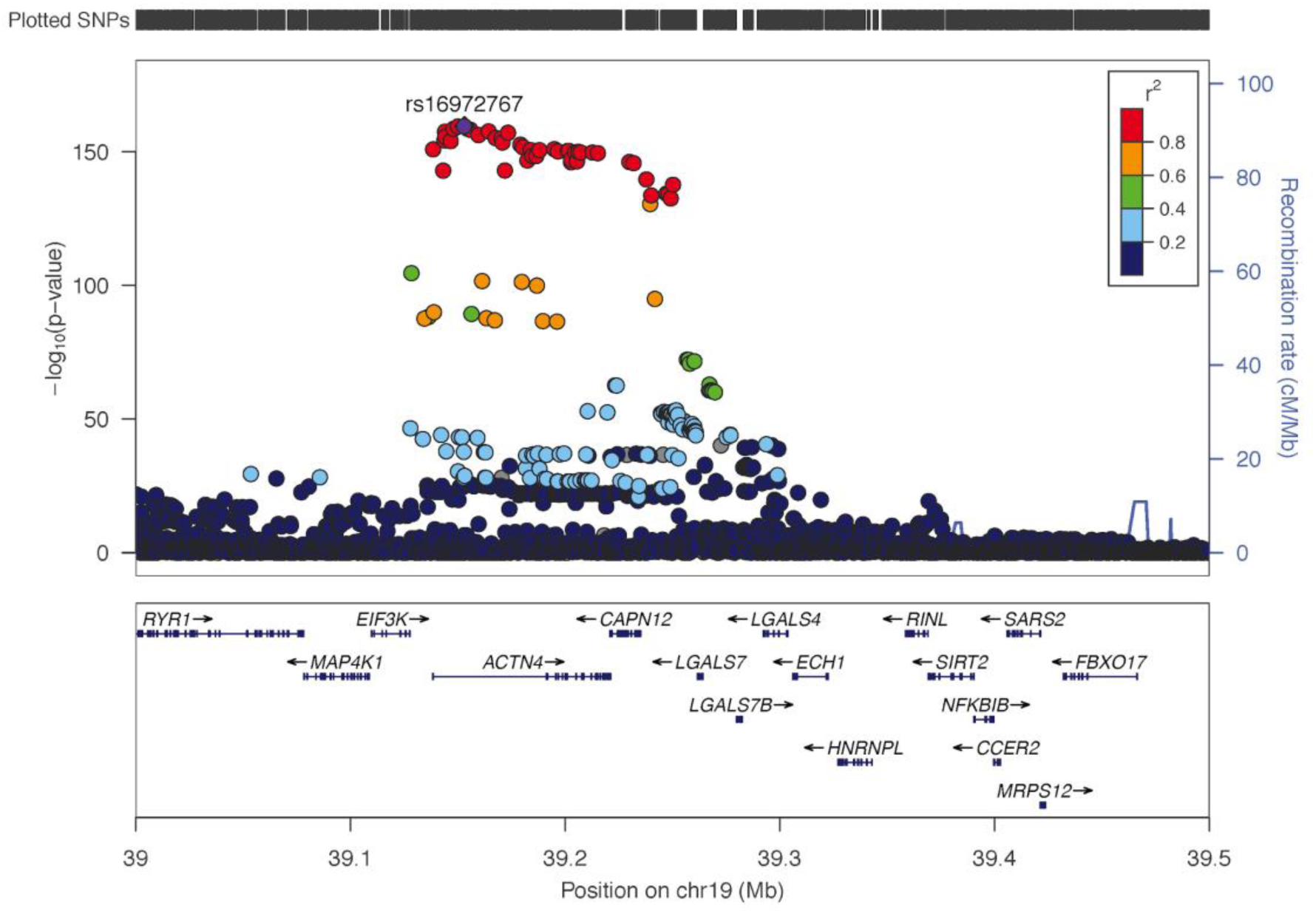
LocusZoom of known associations with rs1808382. Although we did not recover the exact rsid variant, we report a number of exonic variants (more likely to be causative) in extremely strong LD, which represent our strongest signal: in particular, we recovered a variant in perfect LD, rs16972767, which is an intron variant for ACTN4, the gene on which rs1808382 was reported as having potential direct regulatory effects[4]. rs16972767 is reported in the literature as a venular tortuosity hit: in accordance with this, its significance was higher in our venular tortuosity GWAS (-log_10_ p=165) rather than in our arteriolar tortuosity GWAS (-log_10_ p=65).

**Supplemental Figure 12.**
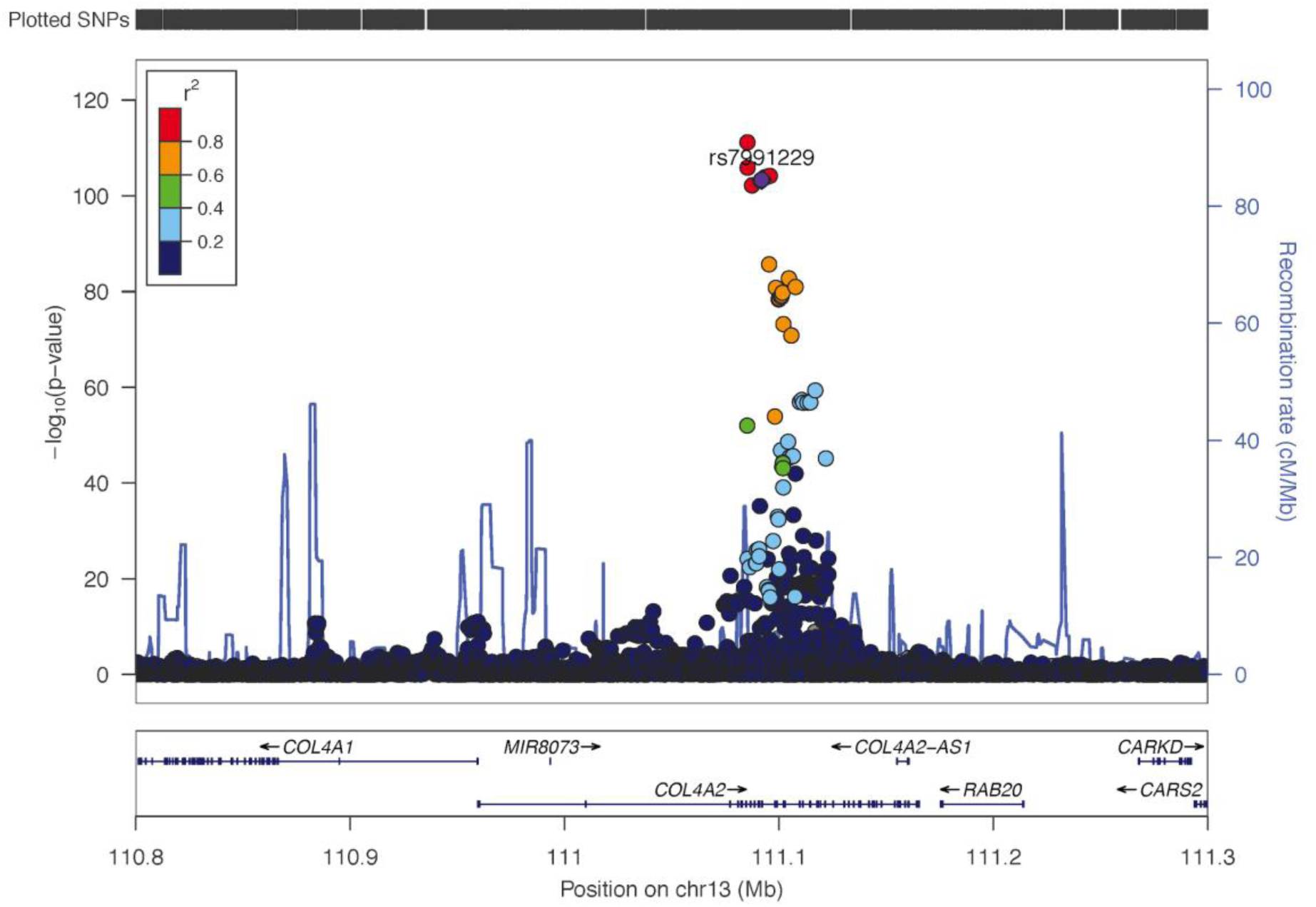
LocusZoom of known associations with rs7991229. This SNP was reported in the literature as an association to arteriolar tortuosity: indeed, its significance was substantially higher in our artery tortuosity GWAS (-log_10_ p=166) than in our vein tortuosity GWAS (-log_10_ p=8).

**Supplemental Figure 13.**
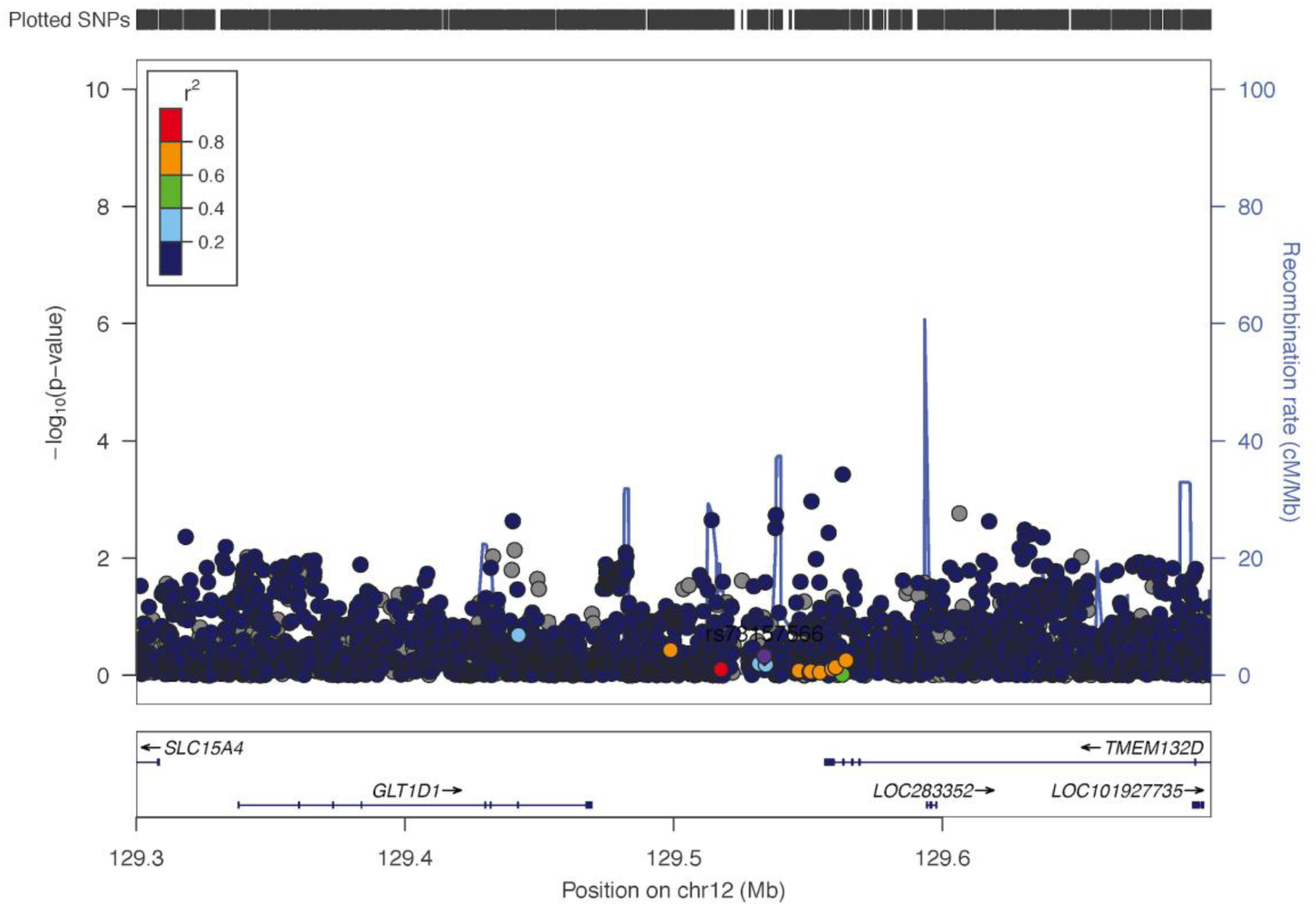
LocusZoom of (controversial) known associations with rs73157566. We did not reproduce it. Given the fact that this association had not reached genome-wide significance in its own replication cohort, we propose this locus should not be considered as associated retinal vessel tortuosity.

### Text 9: Vessel-type comparisons for SNPs, Genes and Pathways

**Supplemental Figure 14.**
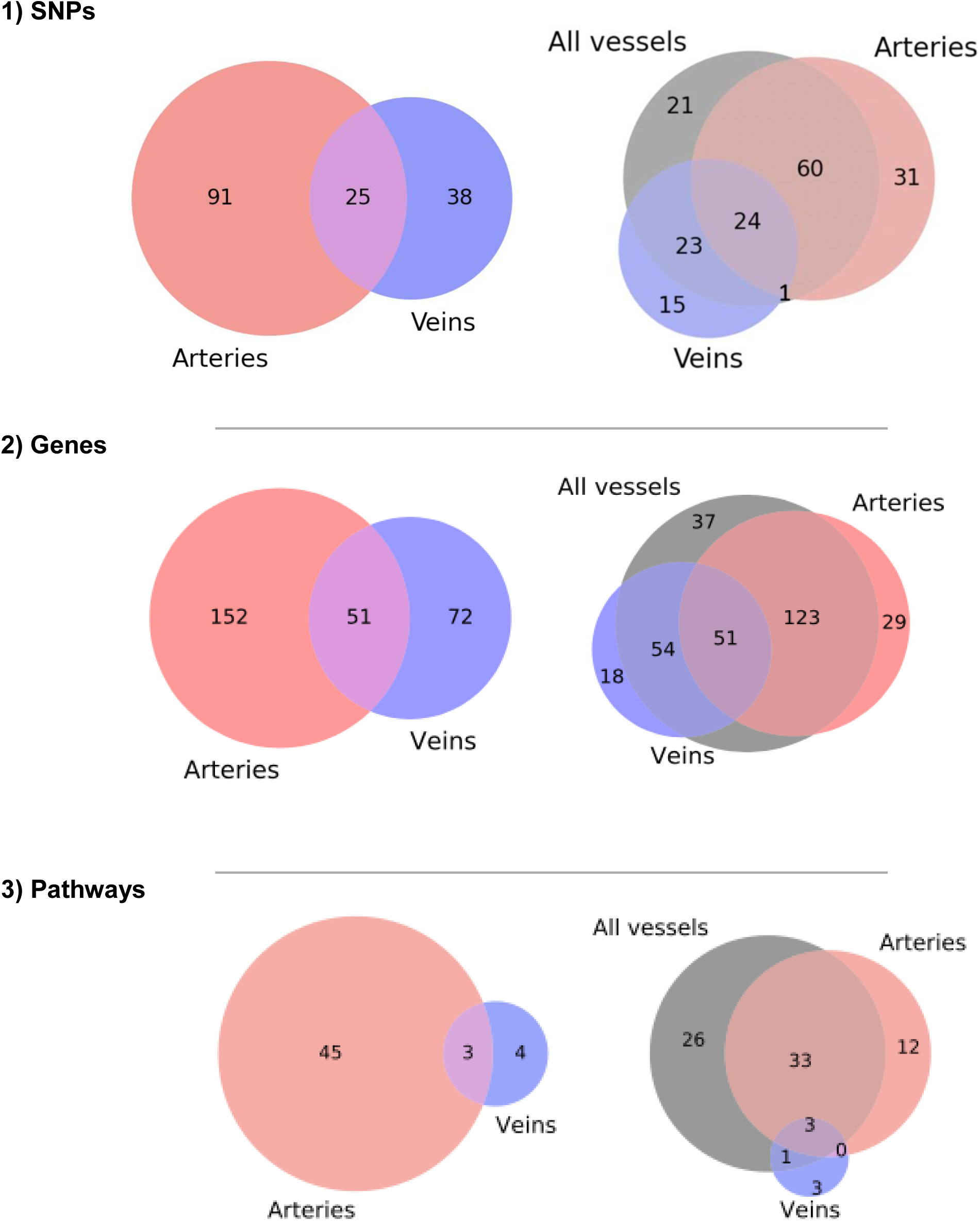
Venn diagrams. Overlap between Bonferroni-thresholded 1) SNPs, 2) genes and 3) pathways across phenotypes. Left: **Hits exclusive to artery GWAS *vs*. vein GWAS:** Venn diagram showing overlap between significant 1) SNPs, 2) genes, 3) pathways in arteries and veins. We find overall more significant SNPS, genes and pathways in arteries, and also more than twice as many artery-exclusive SNPs, genes and pathways than vein-exclusive. Right: **Hits exclusive to artery GWAS *vs*. vein GWAS *vs*. all-vessels (combined-vessel-type GWAS):** Combining arteries and veins into a single “all vessels” phenotype catches many of the SNPs, genes and pathways detected by the artery and vein phenotypes and also provide some news.

### Text 10: Genetic associations with disease and risk

#### Tortuosity variants associated with disease outcome

**Supplemental Table 3.**
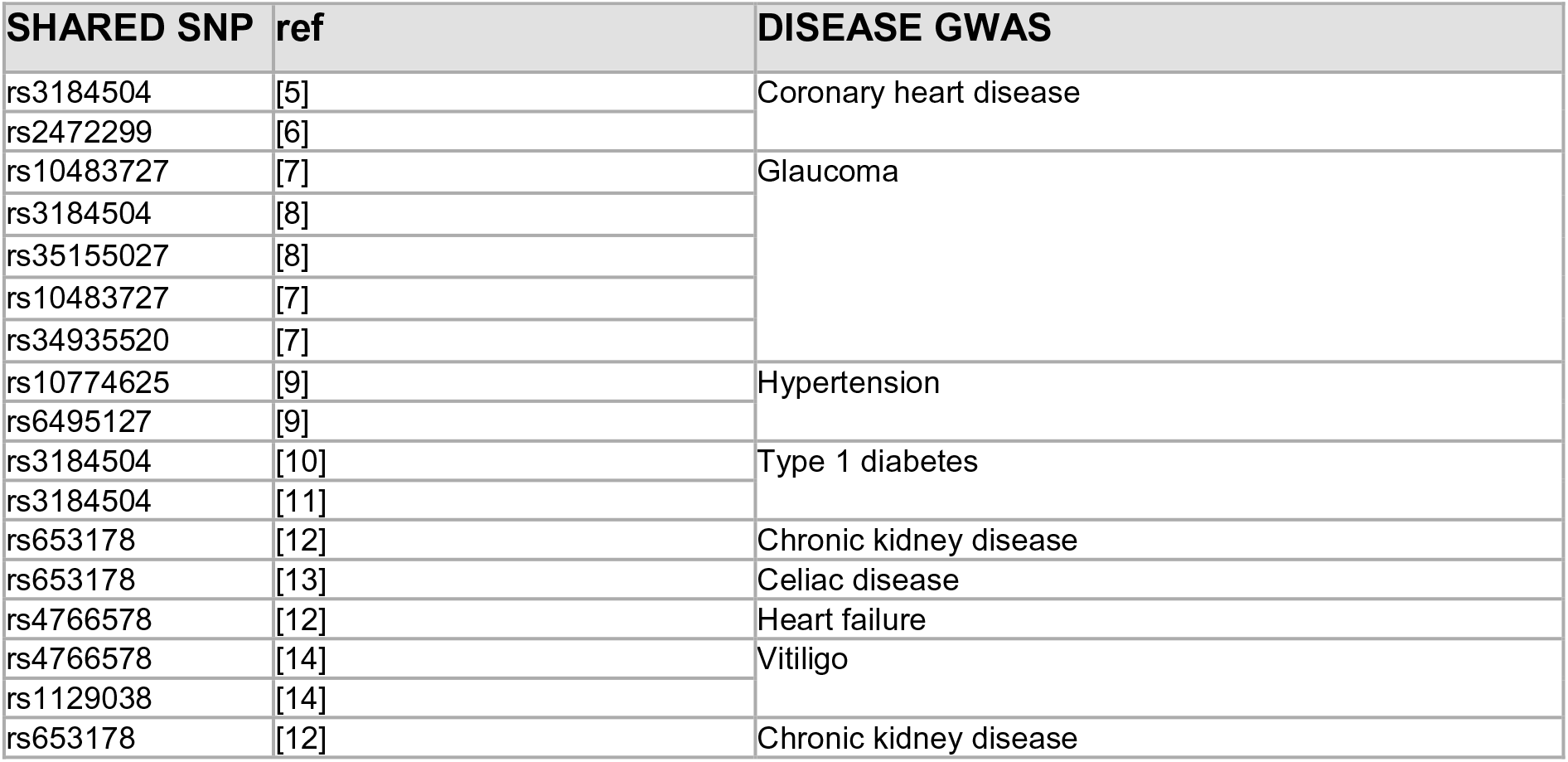
Variants associated with disease. List of variants influencing both tortuosity and disease outcome without statistically significant association to a gene.

**Supplemental Table 4.**
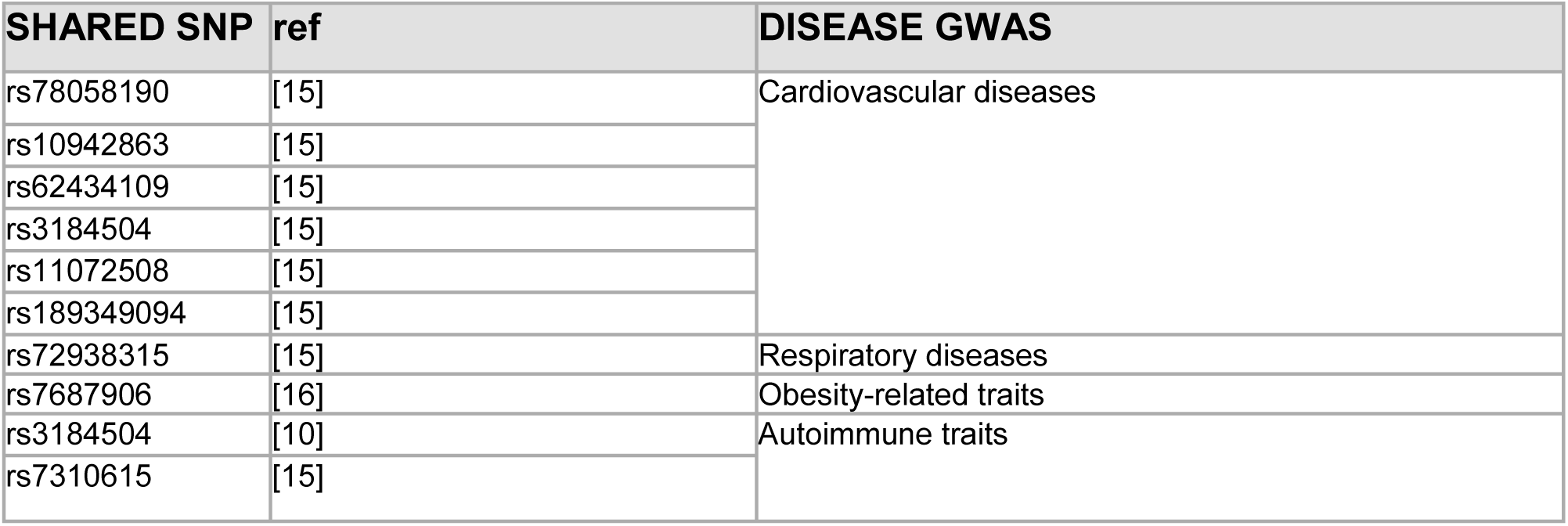
Variants associated with disease (general outcome). List of variants influencing both tortuosity and general non-specific disease outcomes (e.g.,”Obesity-related traits” does not specify the exact disease outcome).

#### Tortuosity variants associated with disease risk factors

**Supplemental Table 5.**
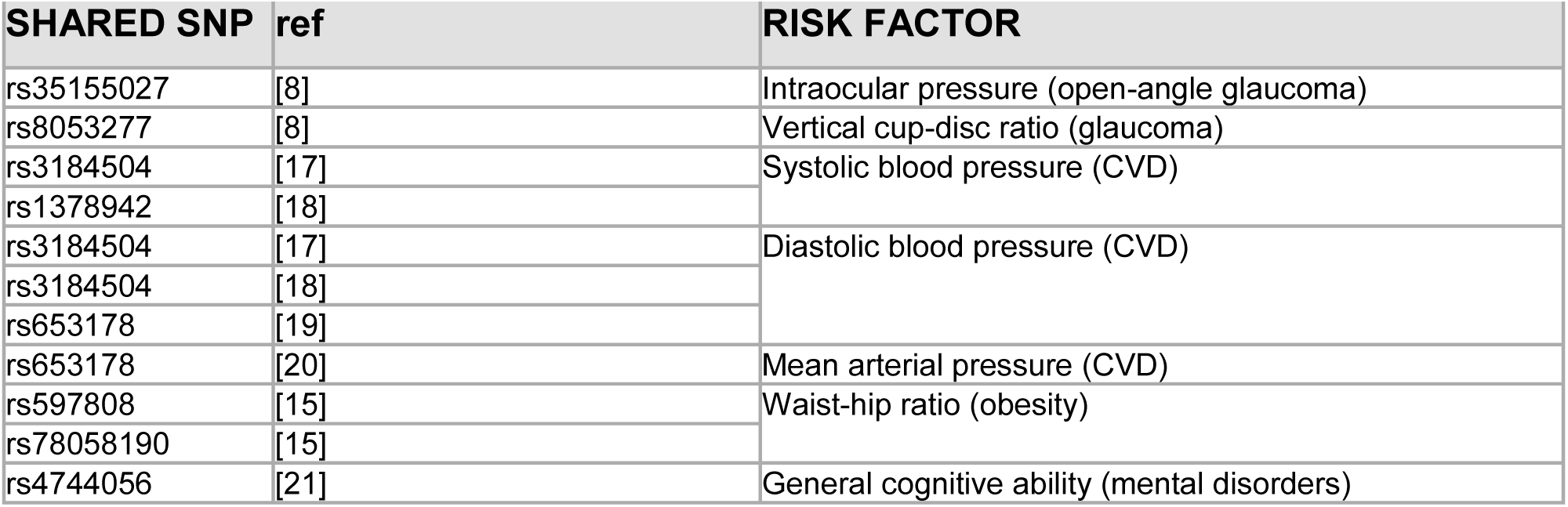
Variants associated with risk of disease. List of variants influencing both tortuosity and disease outcome without statistically significant association to a gene.

### Text 11: Mendelian Randomization

**Supplemental Table 6.** Bi-directional Mendelian Randomization results. Causal estimates are based on the inverse variance weighted (IVW) method. Abbreviations: Body Mass Index (BMI), Coronary artery disease (CAD), Systolic Blood Pressure (SBP), High-density lipoprotein (HDL), Low-density lipoprotein (LDL), total cholesterol (TC) and triglycerides (TG). No multiple hypothesis correction was applied.

| Tortuosity -> Outcome |  |  |  |  | Exposure -> Tortuosity |  |  |  |  |
| --- | --- | --- | --- | --- | --- | --- | --- | --- | --- |
| Exposure | Outcome | Effect | SE | Pvalue | Exposure | Outcome | Effect | SE | Pvalue |
| artery | BMI | 0.019 | 0.015 | 0.200 | BMI | artery | 0.028 | 0.030 | 0.356 |
| mixed |  | 0.008 | 0.015 | 0.581 |  | mixed | -0.011 | 0.029 | 0.715 |
| vein |  | -0.044 | 0.018 | 0.013 |  | vein | -0.047 | 0.027 | 0.078 |
| artery | CAD | -0.053 | 0.041 | 0.200 | CAD | artery | -0.018 | 0.028 | 0.524 |
| mixed |  | -0.061 | 0.038 | 0.109 |  | mixed | -0.008 | 0.024 | 0.748 |
| vein |  | -0.076 | 0.046 | 0.097 |  | vein | -0.002 | 0.016 | 0.924 |
| artery | SBP | -0.200 | 0.362 | 0.581 | SBP | artery | -0.017 | 0.017 | 0.324 |
| mixed |  | 0.033 | 0.344 | 0.924 |  | mixed | -0.015 | 0.015 | 0.309 |
| vein |  | -0.035 | 0.538 | 0.948 |  | vein | -0.010 | 0.009 | 0.252 |
| artery | HDL | 0.017 | 0.015 | 0.236 | HDL | artery | 0.010 | 0.008 | 0.235 |
| mixed |  | 0.014 | 0.015 | 0.351 |  | mixed | 0.011 | 0.009 | 0.193 |
| vein |  | 0.009 | 0.024 | 0.722 |  | vein | 0.008 | 0.009 | 0.383 |
| artery | LDL | -0.001 | 0.016 | 0.940 | LDL | artery | -0.046 | 0.013 | 0.001 |
| mixed |  | 0.006 | 0.017 | 0.709 |  | mixed | -0.043 | 0.013 | 0.001 |
| vein |  | 0.002 | 0.029 | 0.945 |  | vein | -0.031 | 0.013 | 0.018 |
| artery | TC | 0.000 | 0.017 | 0.991 | TC | artery | 0.000 | 0.002 | 0.947 |
| mixed |  | 0.006 | 0.019 | 0.741 |  | mixed | 0.000 | 0.002 | 0.960 |
| vein |  | -0.006 | 0.032 | 0.844 |  | vein | 0.000 | 0.002 | 0.963 |
| artery | TG | -0.006 | 0.013 | 0.656 | TG | artery | -0.038 | 0.018 | 0.038 |
| mixed |  | 0.003 | 0.013 | 0.833 |  | mixed | -0.034 | 0.016 | 0.038 |
| vein |  | -0.008 | 0.022 | 0.720 |  | vein | -0.014 | 0.014 | 0.301 |

### Text 12: ACTN4 and COL4A2 over-expression

**Supplemental Figure 15.**
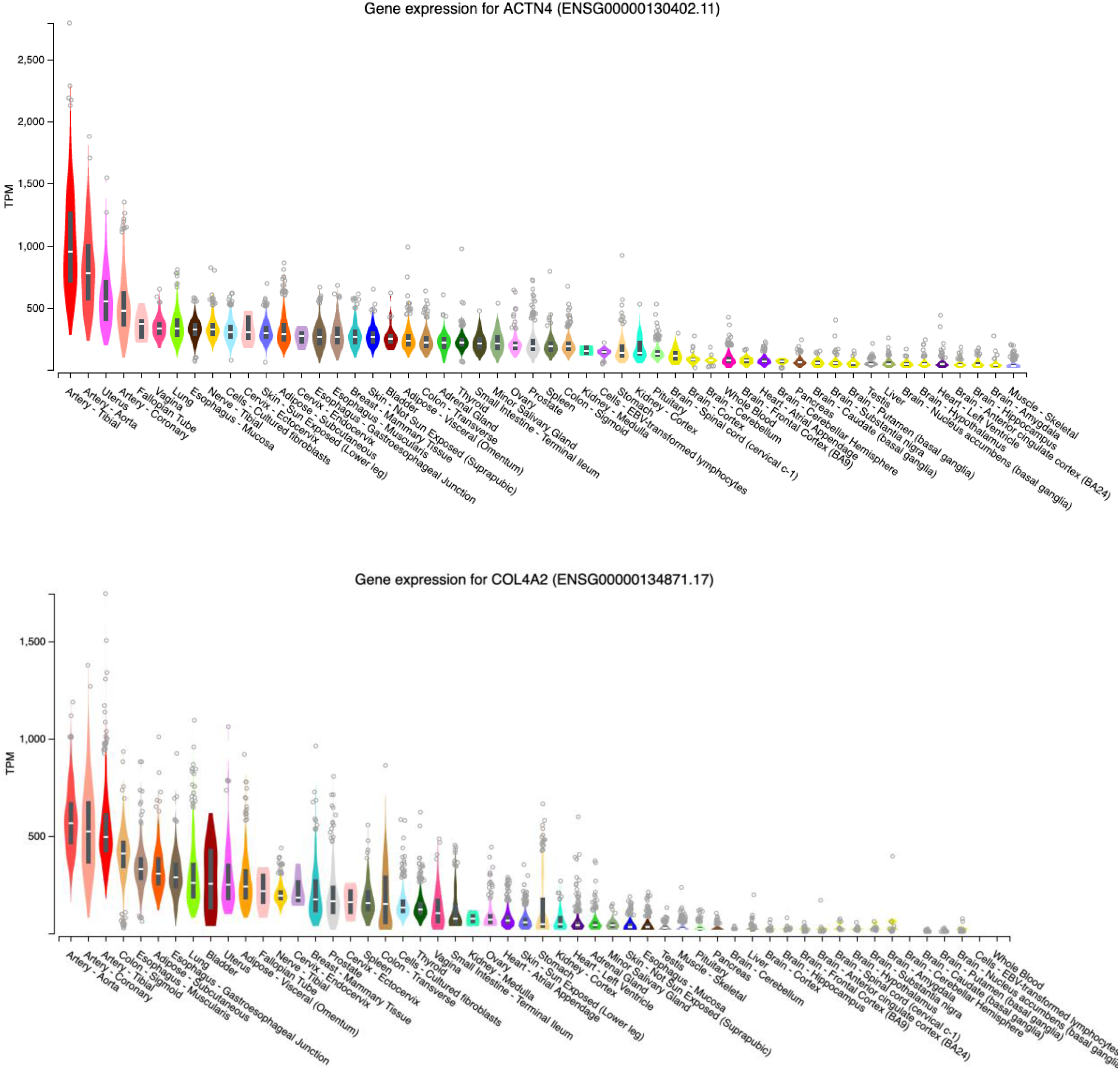
ACTN4 and COL4A2 abundance according to GTEx. The two highly significant genes’ ACTN4 and COL4A2 mRNAs were both found to be highly abundant in blood vessels.

### Text 13: Full gen set enrichment results

**Supplemental Figure 16.**
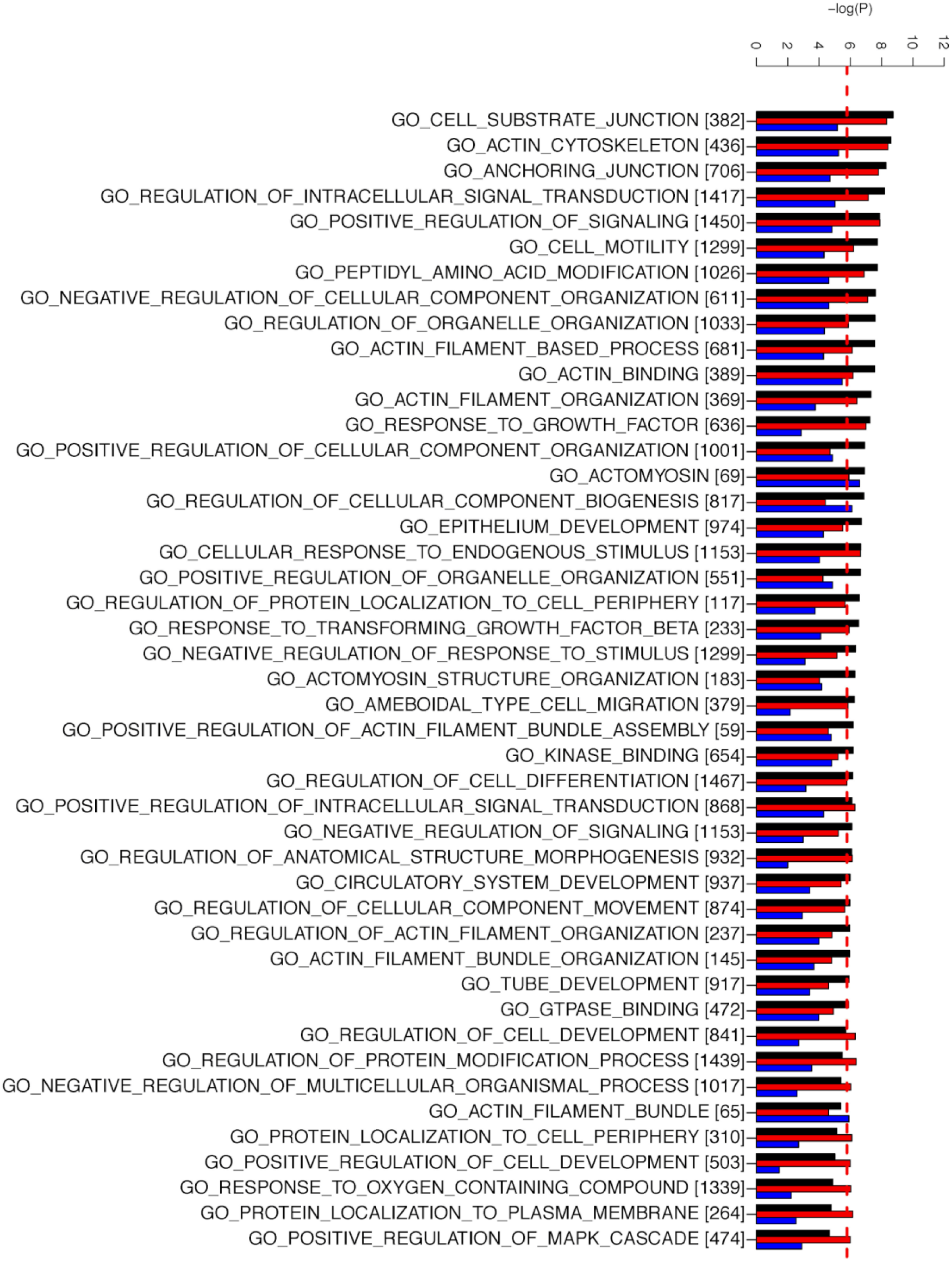
Extended version of Figure 5 from the main text (GO terms). Extended version where all enriched GO terms are shown (irrespective of them being very general or redundant). Additionally, label names are printed out in full. The number of genes in each set is indicated in squared brackets.

**Supplemental Figure 17.**
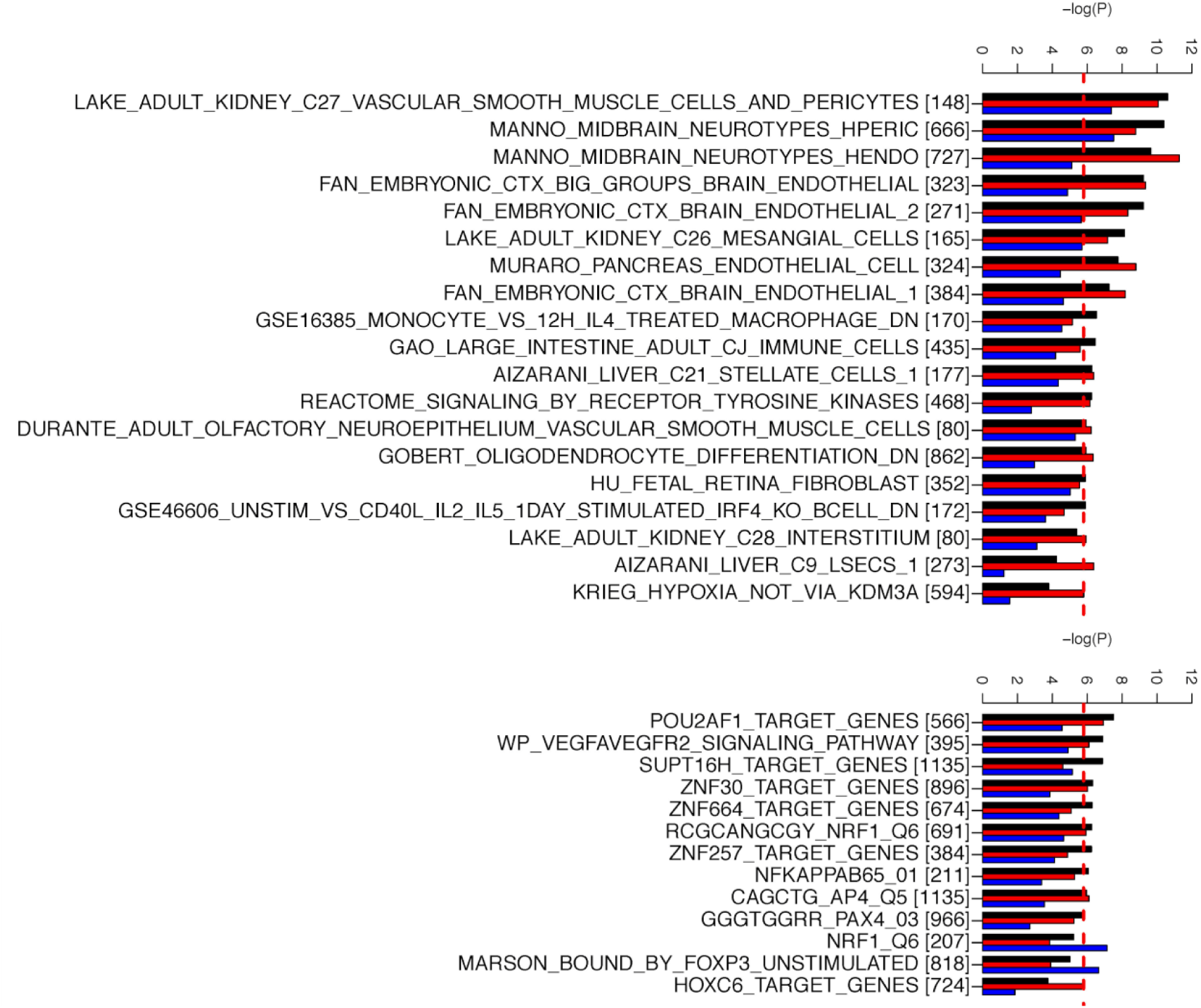
Extended version of Figure 5 from the main text (pathways). Extended version where all enriched pathways are shown (irrespective of them being very general or redundant). Additionally, label names are printed out in full. The number of genes in each set is indicated in squared brackets.

**Supplemental Figure 18.**
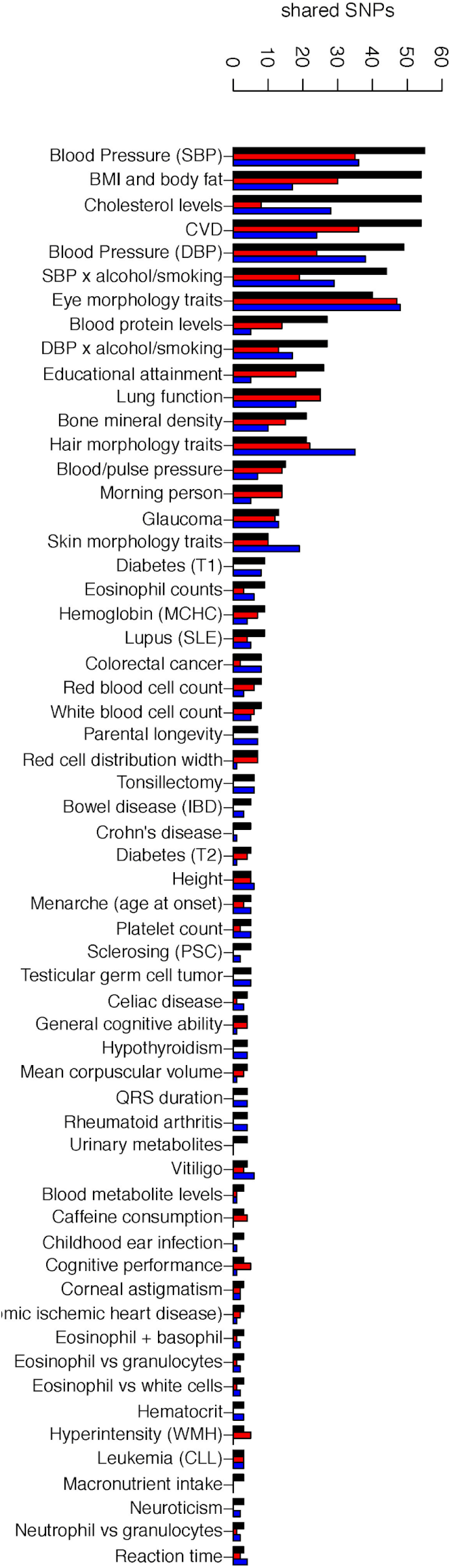
Extended version of Figure 4 from the main text. Extended version where all traits with at least 3 shared associations are included. For full data, refer to Supplemental Dataset 3.

#### List of label replacements

Supplemental Figure 18 was generated based on data in Supplemental Dataset 3. The following replacements were applied to the column “Trait” to homogenize or shorten some labels.

“Blood pressure”<-”Blood/pulse pressure”

“Pulse pressure”<-”Blood/pulse pressure”

“Systolic blood pressure”<-”Blood Pressure (SBP)”

“Mean arterial pressure”<-”Blood Pressure (SBP)”

“Diastolic blood pressure”<-”Blood Pressure (DBP)”

“Blood pressure traits (multi-trait analysis)”<-”Blood Pressure (DBP)”

“Body mass index”<-”BMI and body fat”

“Body fat percentage”<-”BMI and body fat”

“Fat-free mass”<-”BMI and body fat”

“Body mass index (joint analysis main effects and physical activity interaction)”<-”BMI and body fat”

“BMI and body fat (joint analysis main effects and physical activity interaction)”<-”BMI and body fat”

“Waist-to-hip ratio adjusted for BMI (additive genetic model)”<-”BMI and body fat”

“Waist-hip ratio”<-”BMI and body fat”

“Hip circumference adjusted for BMI”<-”BMI and body fat”

“Obesity-related traits”<-”BMI and body fat”

“Triglycerides”<-”Cholesterol levels”

“Cholesterol, total”<-”Cholesterol levels”

“LDL cholesterol”<-”Cholesterol levels”

“High density lipoprotein cholesterol levels”<-”Cholesterol levels”

“Low density lipoprotein cholesterol levels”<-”Cholesterol levels”

“Total cholesterol levels”<-”Cholesterol levels”

“HDL cholesterol”<-”Cholesterol levels”

“C-reactive protein levels or LDL-cholesterol levels (pleiotropy)”<-”Cholesterol levels”

“Coronary artery disease (myocardial infarction, percutaneous transluminal coronary angioplasty, coronary artery bypass grafting, angina or chronic ischemic heart disease)”<-”CVD”

“Myocardial infarction (early onset)”<-”CVD”

“Coronary artery disease or ischemic stroke”<-”CVD”

“Coronary artery disease or large artery stroke”<-”CVD”

“Cardiovascular disease”<-”CVD”

“Ischemic stroke”<-”CVD” “Stroke”<-”CVD”

“Stroke (large artery atherosclerosis)”<-”CVD”

“Stroke (small-vessel)”<-”CVD”

“Myocardial infarction”<-”CVD”

“Ischemic stroke (small-vessel)”<-”CVD”

“Coronary artery disease”<-”CVD”

“Coronary heart disease”<-”CVD”

“CVD (small-vessel)”<-”CVD”

“Type 1 diabetes”<-”Diabetes (T1)”

“Latent autoimmune diabetes vs. type 2 diabetes”<-”Diabetes (T1)”

“Latent autoimmune diabetes vs. type 2 diabetes”<-”Diabetes (T2)”

“Type 2 diabetes”<-”Diabetes (T2)”

“Intraocular pressure”<-”Eye morphology traits”

“Macular thickness”<-”Eye morphology traits”

“Vertical cup-disc ratio”<-”Eye morphology traits”

“Optic cup area”<-”Eye morphology traits”

“Optic disc area”<-”Eye morphology traits”

“Eye color traits”<-”Eye morphology traits”

“Eye color”<-”Eye morphology traits”

“Eye color (brightness)”<-”Eye morphology traits”

“Eye color (hue)”<-”Eye morphology traits”

“Retinal vascular caliber”<-”Eye morphology traits”

“Eye color (saturation)”<-”Eye morphology traits”

“Blue vs. green eyes”<-”Eye morphology traits”

“Blue vs. brown eyes”<-”Eye morphology traits”

“Optic disc parameters”<-”Eye morphology traits”

“Optic disc parameters”<-”Hair morphology traits”

“Hair color”<-”Hair morphology traits”

“Hair morphology traits”<-”Hair morphology traits”

“Blond vs. brown/black hair color”<-”Hair morphology traits”

“Black vs. blond hair color”<-”Hair morphology traits”

“Black vs. red hair color”<-”Hair morphology traits”

“Blond vs. brown hair color”<-”Hair morphology traits”

“Brown vs. black hair color”<-”Hair morphology traits”

“Red vs. brown/black hair color”<-”Hair morphology traits”

“Skin pigmentation traits”<-”Skin morphology traits”

“Skin pigmentation”<-”Skin morphology traits”

“Perceived skin darkness”<-”Skin morphology traits”

“Glaucoma (primary open-angle)”<-”Glaucoma”

“Glaucoma (high intraocular pressure)”<-”Glaucoma”

“Lung function (FEV1/FVC)”<-”Lung function”

“Lung function (FEV1)”<-”Lung function”

“Post bronchodilator FEV1/FVC ratio”<-”Lung function”

“FEV1”<-”Lung function”

“Peak expiratory flow”<-”Lung function”

“Lung function (FVC)”<-”Lung function”

“Lung function (Lung function)”<-”Lung function”

“Mean arterial pressure x alcohol consumption (light vs heavy) interaction (2df test)”<-”SBP x alcohol/smoking”

“Mean arterial pressure x alcohol consumption interaction (2df test)”<-”SBP x alcohol/smoking”

“Blood Pressure (SBP) x alcohol consumption interaction (2df test)”<-”SBP x alcohol/smoking”

“Systolic blood pressure (alcohol consumption interaction)”<-”SBP x alcohol/smoking”

“Blood Pressure (DBP) x alcohol consumption interaction (2df test)”<-”DBP x alcohol/smoking”

“Diastolic blood pressure x smoking status (current vs non-current) interaction (2df test)”<-”DBP x alcohol/smoking”

“Diastolic blood pressure x smoking status (ever vs never) interaction (2df test)”<-”DBP x alcohol/smoking”

“Blood Pressure (SBP) x alcohol consumption (light vs heavy) interaction (2df test)”<-”SBP x alcohol/smoking”

“Blood Pressure (DBP) x alcohol consumption (light vs heavy) interaction (2df test)”<-”DBP x alcohol/smoking”

“Blood Pressure (SBP) (cigarette smoking interaction)”<-”SBP x alcohol/smoking”

“Blood Pressure (DBP) (cigarette smoking interaction)”<-”DBP x alcohol/smoking”

“Systolic blood pressure x alcohol consumption interaction (2df test)”<-”SBP x alcohol/smoking”

“Diastolic blood pressure x alcohol consumption interaction (2df test)”<-”DBP x alcohol/smoking”

“Systolic blood pressure x alcohol consumption (light vs heavy) interaction (2df test)”<-”SBP x alcohol/smoking”

“Diastolic blood pressure x alcohol consumption (light vs heavy) interaction (2df test)”<-”DBP x alcohol/smoking”

“Systolic blood pressure (cigarette smoking interaction)”<-”SBP x alcohol/smoking”

“Diastolic blood pressure (cigarette smoking interaction)”<-”DBP x alcohol/smoking”

“Educational attainment (MTAG)”<-”Educational attainment”

“Highest math class taken (MTAG)”<-”Educational attainment”

“Cognitive performance (MTAG)”<-”Cognitive performance”

“Parental longevity (combined parental attained age, Martingale residuals)”<-”Parental longevity”

“Educational attainment (years of education)”<-”Educational attainment”

“Alcohol consumption (drinks per week)”<-”Alcohol (drinks/week)”

“Mean corpuscular hemoglobin”<-”Hemoglobin (MCHC)”

“Mean corpuscular hemoglobin concentration”<-”Hemoglobin (MCHC)”

“Systemic lupus erythematosus”<-”Lupus (SLE)”

“Morning vs. evening chronotype”<-”Morning person”

“Chronotype”<-”Morning person”

“Primary sclerosing cholangitis”<-”Sclerosing (PSC)”

“Urinary metabolites (H-NMR features)”<-”Urinary metabolites”

“Heel bone mineral density”<-”Bone mineral density”

“Chronic lymphocytic leukemia”<-”Leukemia (CLL)”

“Dietary macronutrient intake”<-”Macronutrient intake”

“Eosinophil percentage of granulocytes”<-”Eosinophil vs granulocytes”

“Eosinophil percentage of white cells”<-”Eosinophil vs white cells”

“Neutrophil percentage of granulocytes”<-”Neutrophil vs granulocytes”

“Sum eosinophil basophil counts”<-”Eosinophil + basophil”

“White matter hyperintensity burden”<-”Hyperintensity (WMH)”

“Inflammatory bowel disease”<-”Bowel disease (IBD)

